# Transcriptome-wide and Stratified Genomic Structural Equation Modeling Identify Neurobiological Pathways Underlying General and Specific Cognitive Functions

**DOI:** 10.1101/2021.04.30.21256409

**Authors:** Andrew D. Grotzinger, Javier de la Fuente, Gail Davies, Michel G. Nivard, Elliot M. Tucker-Drob

## Abstract

Spearman’s observation in 1904 that distinct cognitive functions—such as reasoning, processing speed, and episodic memory—are positively intercorrelated has given rise to over a century of speculation and investigation into their common and domain-specific mechanisms of variation. Here we develop and validate Transcriptome-wide Structural Equation Modeling (T-SEM), a novel method for studying the effects of tissue-specific gene expression within multivariate space. We apply T-SEM to investigate the shared and unique functional genomic characteristics of seven, distinct cognitive traits (*N* = 11,263–331,679). We identify 184 genes associated with general cognitive function (*g*), including 10 novel genes not identified in univariate analysis for the individual cognitive traits. We go on to apply Stratified Genomic SEM to identify enrichment for *g* within 29 functional genomic categories. This includes categories indexing the intersection of protein-truncating variant intolerant (PI) genes and specific neuronal cell types, which we also find to be enriched for the genetic covariance between *g* and a psychotic disorders factor.

---

The finding that many diverse cognitive functions are positively intercorrelated has been described as “arguably the most replicated result in all psychology,”^1^ and is the basis for the well-known concept of general cognitive ability, *g*. Despite over 100 years of debate regarding the nature and mechanisms of *g*, empirical work into neurobiological mechanisms of *g* have primarily focused on gross anatomical structures derived using brain imaging.^2^ Functional genomic approaches provide powerful insights into the molecular biology of trait variation, but because they have primarily been developed for analysis of one phenotype at a time, have had limited application to the study of constellations of correlated traits.

Transcriptome-Wide Association Studies (TWAS) combine discoveries made regarding the genetic regulation of tissue-specific gene expression to identify biological pathways underlying GWAS phenotypes. T-SEM expands TWAS to the multivariate space within the general Genomic Structural Equation Modelling (Genomic SEM) framework^3^ to allow researchers to examine how tissue-specific expression of individual genes operates within a multivariate system of heritable phenotypes. For example, T-SEM can estimate relationships between gene expression and a general factor, along with a Q_Gene_ statistic that quantifies data that deviate from the hypothesis that gene expression affects the traits strictly via the factor. We demonstrate via simulation that T-SEM is well-calibrated, with the power to detect gene expression effects on a common factor and for Q_Gene_ increasing and decreasing, respectively, as the population generating gene expression effects shift closer to the pattern of relationships implied by the common factor (**Method**; Supplementary Figures 2-5; Supplementary Table 2).

We applied T-SEM to GWAS summary statistics for seven cognitive traits (Table S1) from UK Biobank (UKB): trail-making tests-B, tower rearranging, verbal numerical reasoning (VNR), symbol digit substitution, memory pairs-matching test, matrix pattern recognition, and reaction time (RT). We employ the same common factor model reported in de la Fuente et al.,^4^ who identified a general dimension of shared genetic liability accounting for an average of 58.4% of the genetic variance in these same cognitive traits, which they termed genetic *g* (Supplementary Figure 1). These summary statistics were paired with cis gene expression quantitative trait locus (cis-eQTL) reference panel data for 13 brain-based tissue types from GTEx^5^ and the two dorsolateral prefrontal cortex (dlPFC) datasets from the Common Mind Consortium (CMC)^6^ to produce univariate TWAS estimates in FUSION^7^ for 52,849 genes across tissues, which were then used as input for T-SEM.

T-SEM analyses revealed 218 genes whose inferred expression was significantly associated with *g* at a Bonferroni corrected threshold (Figure 1; Table 1; Supplementary Table 3). Of these 218 hits, 184 were independent from Q_Gene_ hits and may, therefore, be considered as operating directly on *g*. As many genes are expressed across multiple reference tissues, these 184 hits ultimately reflect 76 unique genes, including 10 novel genes that were not significant for any of the univariate TWAS analyses in any tissue type. Gene co-expression analyses using these 76 unique gene IDs as input revealed a total of 59 significant gene sets across three primary clusters (Supplementary Table 5; Supplementary Figure 8). This included a number of gene sets implicated in transfer RNA (tRNA), neuron-specific chromatin regulatory BAF subunits (nBAFs), and G-protein coupled receptors, all of which have been associated with general neural development, cognitive function, and neurodevelopmental disorder risk.^8-10^

**Figure 1.**
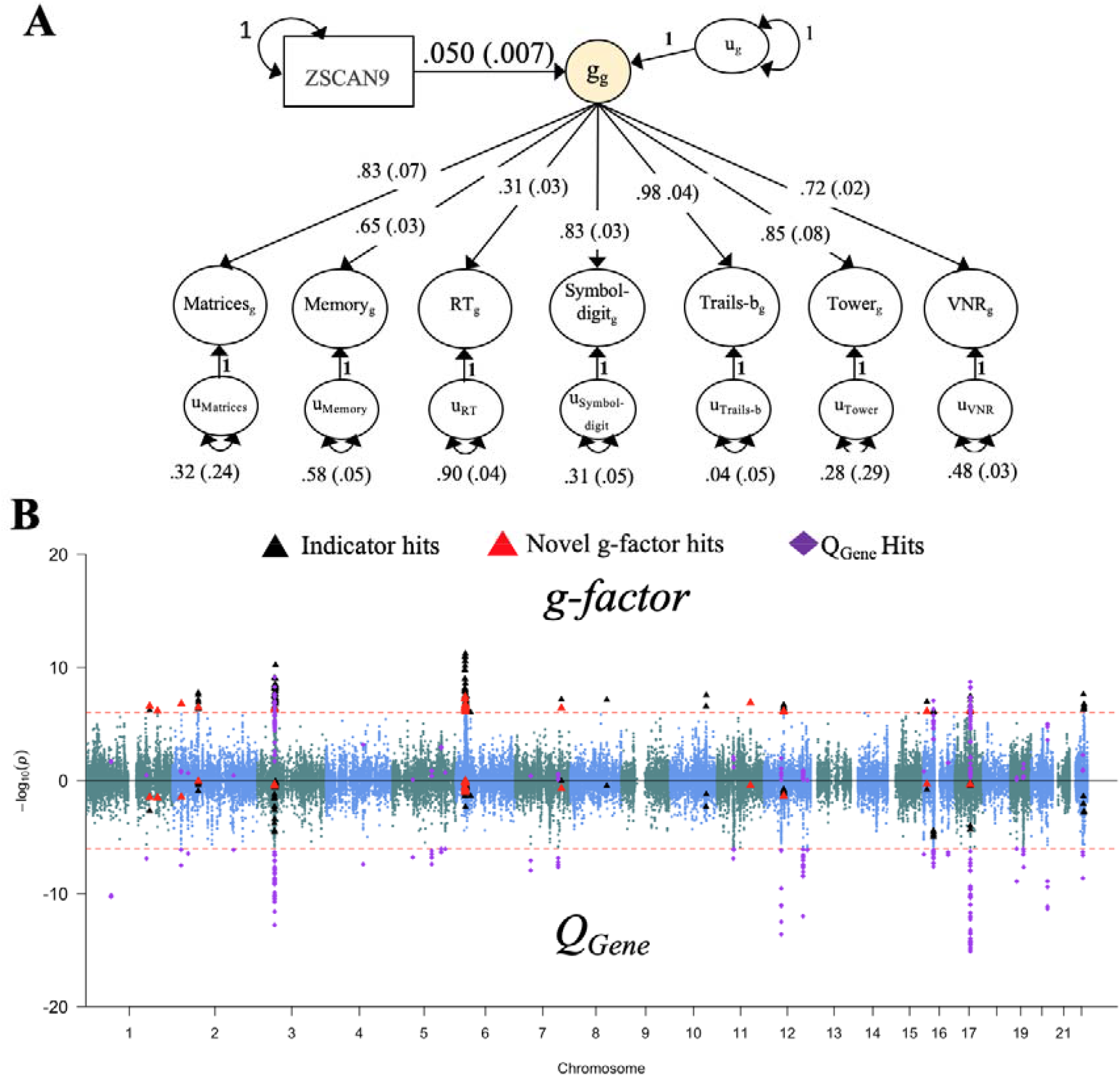
*g*-factor T-SEM. *Panel A* depicts the results from the top gene, ZSCAN9 in the cerebellum, predicting the *g-*factor estimated using T-SEM. Results are standardized with respect to the seven cognitive outcomes, while the variance in the *g-*factor reflects 1 + the variance explained by the individual gene. Standard errors are shown in parentheses. *Panel B* depicts the Miami plot for *g-*factor T-SEM results. The top half of the plots depict the −log10(p) values for TWAS effects on the g-factor; the bottom half depicts the log10(p) values for the g-factor Q_Gene_ effects. The red dashed line marks the threshold for TWAS significance using a Bonferroni corrected threshold for 52,849 tests. Black triangles denote g-factor TWAS hits that overlapped with hits for one of the univariate cognitive indicators. Larger red triangles denote novel TWAS g-factor hits that did not overlap with the univariate indicators or hits for Q_Gene._ Purple diamonds denote Q_Gene_ hits.

**Table 1.** Multivariate TWAS Results.

| TWAS Target | All Tissues |  |  |  | Unique Gene IDs |  |  |
| --- | --- | --- | --- | --- | --- | --- | --- |
|  |  | Hits | Shared Hits | Unique hits | Hits | Shared Hits | Unique hits |
| | Mean $\chi^2(1)$ | (Q<sub>Gene</sub> hits) | (Q<sub>Gene</sub> hits) | (Q<sub>Gene</sub> hits) | (Q<sub>Gene</sub> hits) | (Q<sub>Gene</sub> hits) | (Q<sub>Gene</sub> hits) |
| *g-factor* | 2.13 | 218 (34) | 184 (34) | 34 (0) | 88 (12) | 78 (12) | 10 (0) |
| *QGene* | 1.91 | 156 | 133 | 23 | 62 | 41 | 21 |
| *Matrix* | 1.09 | 1 (0) | 0 (0) | 1 (0) | 1 (0) | 0 (0) | 1 (0) |
| *Memory* | 1.50 | 17 (0) | 1 (0) | 16 (0) | 12 (0) | 1 (0) | 11 (0) |
| *Reaction Time* | 1.95 | 119 (70) | 19 (16) | 100 (54) | 49 (20) | 5 (4) | 44 (16) |
| *Symbol Digit* | 1.39 | 3 (0) | 2 (0) | 1 (0) | 3 (0) | 2 (0) | 1 (0) |
| *TMTB* | 1.46 | 25 (3) | 22 (3) | 3 (0) | 16 (2) | 15 (2) | 1 (0) |
| *Tower* | 1.05 | 0 (0) | 0 (0) | 0 (0) | 0 (0) | 0 (0) | 0 (0) |
| *VNR* | 2.56 | 507 (67) | 162 (18) | 345 (49) | 183 (24) | 70 (9) | 113 (15) |

*Note.* Values in parentheses indicate whether any of the TWAS hits were also hits for $Q_{\text{Gene}}$ within the same tissue type. $Q_{\text{Gene}}$ indexes whether a particular gene is unlikely to operate through the identified $g$ -factor structure, as will often be the case when a gene effect is highly specific to an individual trait. To facilitate comparison across TWAS targets, mean $\chi^2$ values reported in each row were converted to $\chi^2(1)$ statistics before taking their means. Unique gene IDs were defined as those genes that were significant across any tissue. TMTB = trail making test-b; VNR = verbal numerical reasoning.

Using these 184 T-SEM hits for *g* as input, joint analyses revealed 29 conditionally significant genes across 18 loci. These 18 loci explained, on average, 81.1% of the variance of nearby GWAS estimates (range = 58% - 100%; Supplementary Table 4; Supplementary Figure 6 for regional association plots). In addition, the conditional significance of the top SNP within these loci dropped substantially from an average *p*-value of 1.84E-6 to .153, with none of the previous top SNPs within these loci remaining nominally significant. This indicates that inferred gene expression patterns generally account for a large portion of observed GWAS effects on *g*. Bayesian colocalization analysis were additionally used to examine shared causal variants between hits for gene expression and the *g*-factor GWAS (**Method**). These results revealed that a majority (58.2%) of the 184 *g* T-SEM hits were supported by a model of colocalized gene expression and GWAS associations (mean posterior probability = .568; Supplementary Table 4), with a smaller subset (19.6%; mean posterior probability = .223) likely characterized by independent associations. Conservative permutation tests were also used to produce empirically derived univariate TWAS *p*-values used as input for T-SEM (Supplementary Table 4; **Method**). As is expected for the permutation test, these results indicated an overall attenuation in signal, but largely supported the current findings.

For Q_Gene_, we identified 156 hits reflecting 62 unique genes (Supplementary Table 6). These hits reflect genes whose inferred expression is associated with the cognitive traits according to patterns that are inconsistent with their operation on genetic *g*. Similar to *g*-factor hits, Bayesian colocalization analyses supported a model of colocalized associations for the majority of Q_Gene_ hits (51.3%; mean posterior probability = .506), while 22.4% of hits likely indexed independent functional and GWAS hits (mean posterior probability = .242). Among the most significant Q_Gene_ hits were 4 unique genes (NSFP1, NSF, ARL17B, LRRC37A) in the 17q21.31 locus, a region linked to frontotemporal dementia^11^ and autism.^12^ This collection of Q_Gene_ hits consistently evinced a much stronger association with RT relative to the remaining, six cognitive phenotypes (Supplementary Table 6; Supplementary Figure 9), indicating this region is more relevant to cognitive speed than overall genetic *g*. Within particular tissues, we observed a higher relative mean χ^2^ and density of hits for the *g*-factor relative to Q_Gene,_ including in the frontal cortex and hippocampus (Supplementary Table 7). Consistent with conceptualizations of the cerebellum as particularly relevant to broad cognitive function,^13^ this specific tissue showed the largest *g*-factor to Q_Gene_ χ^2^ ratio (Supplementary Figure 10). We go on to identify additional biological pathways underlying genetic *g* using Stratified Genomic SEM.

Stratified Genomic SEM is a recently developed method that can be used to examine enrichment of any model parameter estimated in Genomic SEM, thereby allowing for examining functional enrichment within a multivariate space.^14^ This facilitates identifying categories for which pleiotropic variation, as separable from trait-specific genetic variation, is enriched. Using QC and analytic procedures outlined in the **Method** section, enrichment analyses were based on 155 binary annotations (see **Supplementary Method** for enrichment results in standardized space). We observed 28 annotations that were significant for *g* at a Bonferroni corrected threshold for 155 tests (Figure 2; Supplementary Table 8; Supplementary Figure 12). In line with prior functional findings for intelligence^15^ and general cognitive function,^16^ which were based on univariate methods, we observe significant enrichment in conserved regions and the H3K9ac promoter. This indicates that these previous discoveries generalize to the genetic architecture that is shared across multiple domains of cognitive function. We additionally observe a number of significantly enriched annotations for the H3K27ac promoter across different brain regions (e.g., dlPFC, middle hippocampus). Of these 28 significant annotations, four reflected the intersection of PI genes and the GABAergic and excitatory hippocampal and prefrontal cortex neuronal subtypes (Figure 2).

**Figure 2.**
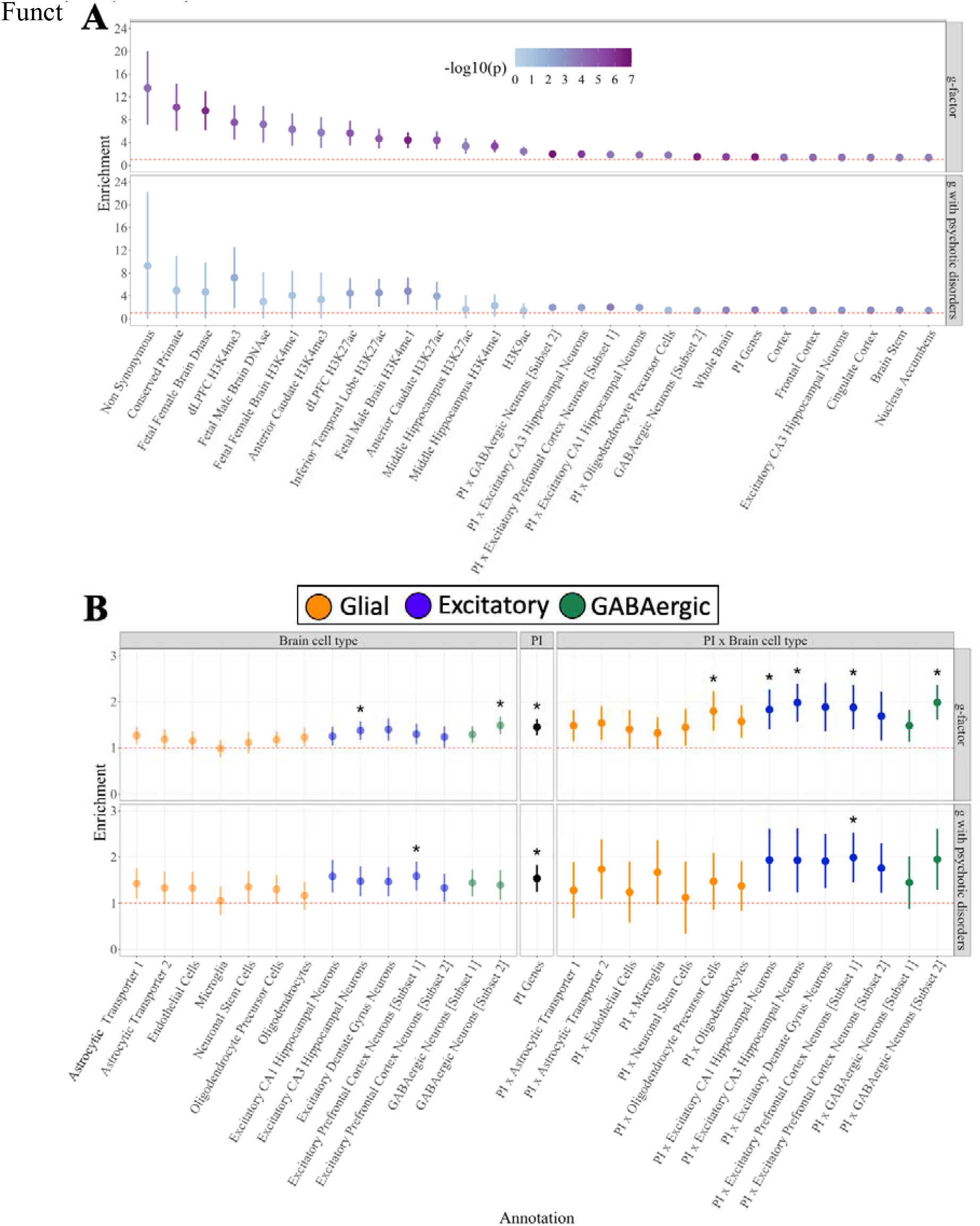
Multivariate Enrichment. *Panel A* displays the 28 annotations for the *g-*factor that were significant at Bonferroni corrected threshold for 155 tests. Dots are ordered according to the size of the enrichment point estimate for the *g*-factor and shaded to reflect level of significance. *Panel B* depicts the PI x brain cell annotations and orders the estimates by brain cell type, PI genes, and PI x brain cell type interactions. Glial cells depicted in orange, excitatory cells in blue, and GABAergic cells in green. Dots that were significant at a Bonferroni corrected threshold for 155 tests are depicted with a *. In both panels A and B, the red dashed line reflects the null (enrichment = 1), error bars depict 95% CIs, and the top half within each panel depicts enrichment estimates for the *g*-factor and the bottom half enrichment estimates for the genetic covariance between the *g*-factor and the psychotic disorders factor.

We observed a number of significant enrichment estimates for the residual variance components (Supplementary Table 8; Supplementary Figure 13). These estimates reflect enrichment of trait-specific genetic variation as separable from genetic *g*. As might be expected, significant estimates were identified for the three cognitive traits with the smallest factor loadings, with 38 significant estimates observed for RT, 33 for VNR, five for the memory pairs-matching test, and no significant estimates for the remaining four traits (Supplementary Figure 14). Among these significant residual estimates, it is perhaps most interesting to consider those that also evinced a weak signal for *g*. This included enrichment for the residuals of VNR and RT for the H3K9ac promoter in the dlPFC, which has been previously associated with Alzheimer’s disease,^17^ and enrichment of the FANTOM5 enhancer, an annotation strongly linked to immunological diseases,^18^ for memory pairs-matching.

The PI enrichment findings for *g* were notably similar to a pattern of enrichment recently described for a psychotic disorders factor defined by bipolar disorder and schizophrenia.^14^ As we also observed a sizeable, negative genetic correlation between the *g*-factor and a psychotic disorders factor (*r*_g_ = −.42, *SE* = .04; Supplementary Figure 11), we went on to examine enrichment of the genetic sharing between these factors (Supplementary Table 8). Results revealed significant enrichment of the factor covariance for three annotations: PI genes, excitatory prefrontal cortex neurons, and their intersection (Supplementary Figure 15). These may represent specific biological pathways underlying the well-established association between cognitive impairment and general risk for disorders with psychotic features.^19,20^

Cognitive functions are characterized by positive intercorrelations at both the observed and genetic levels of analysis. We have used multivariate functional genomic methods to identify both general and trait-specific biological mechanisms of variation across seven cognitive functions. Using T-SEM, we identified 76 unique genes whose inferred expression acts generally across all seven cognitive traits. Highlighting the ability of multivariate methods to leverage shared power for novel discovery, this included 10 genes that were not significant for univariate TWAS of any of the individual traits. Q_Gene,_ a measure of heterogeneity, identified an additional 62 unique genes whose expression affects individual cognitive traits not via *g*. This included a set of genes on locus 17q21.31 that appear highly specific to cognitive speed. Applying Stratified Genomic SEM, we identified 28 annotations significantly enriched for *g*, including evolutionarily conserved annotations and promoters in various brain regions. Specific annotations based on genes predominantly expressed in excitatory prefrontal cortex neurons were enriched in their contribution to the genetic covariance between *g* and disorders with psychotic features. These findings collectively offer critical insights into the biology underlying the genetic variation that is shared and distinct across cognitive traits.

## Online Method

### Multivariate TWAS in Genomic SEM: T-SEM

The general goal of TWAS can be summarized as an effort to uncover the association between gene expression and the outcome(s) of interest. Univariate TWAS works by combining information from both gene expression reference panel data and external GWAS datasets to perform summary-based transcriptomic imputation (TI)/TWAS for a given complex trait. Thus, TWAS does not require that gene expression data be directly available for the GWAS outcome. Transcriptome-wide Structural Equation Models (T-SEM) then draw on univariate, summary-based TWAS produced for multiple GWAS phenotypes in order to analyze the relationships across inferred, gene expression-trait associations in a multivariate space. In practice, we specifically utilize the FUSION software^7^ to perform univariate, summary-based TWAS, which imputes the relationship between gene expression and a trait using the linear combination of GWAS Z-statistics and a set of functional weights. For the currently analyses, we use the precomputed functional weights available directly from the FUSION website (http://gusevlab.org/projects/fusion/) from cis gene expression quantitative trait locus (cis-eQTL) reference panels (see **Univariate TWAS** section below for details of our specific data sources). These weights are obtained in FUSION by comparing the performance of five different penalized linear models: best linear unbiased predictor (BLUP), Bayesian sparse linear model (BSLMM), elastic-net regression (eNET), lasso regression (LASSO), and single best eQTL (top1).^7^ For each gene, the weights are used from the model that produces the largest R^2^ between the predicted and observed expression models calculated using five-fold cross-validation.

T-SEM estimation follows the general two-stage approach introduced in the Genomic SEM framework,^3^ with the goal of modeling genetic covariance between various traits, and the genetically imputed expression level of a gene. In Stage 1 of T-SEM, the genetic covariance matrix and associated sampling covariance matrix across multiple traits are estimated via joint analysis of the univariate GWAS summary statistics for each phenotype in the model using multivariable LDSC. This genetic covariance matrix, which we term *S*_*LDSC*_, contains SNP heritabilities on the diagonal and genetic covariances on the off-diagonal. The sampling covariance matrix, which we term *V*_*SLDSC*_, is a symmetric matrix composed of the nonredundant elements in the *S*_*LDSC*_ matrix. The diagonal elements of *V*_*SLDSC*_ are squared *SE*s of the elements in *S*_*LDSC*_. The off-diagonal elements are sampling covariances that index dependencies across estimation errors. These sampling covariances are estimated using a block-jackknife procedure that quantifies the extent to which the sampling distributions of different elements in the *S*_*LDSC*_ matrix covary with one another, as would be expected when there is sample overlap across the included traits. The combination of diagonal and off-diagonal elements is what allows Genomic SEM to produce unbiased SEs in the context of the user-specific structural models, even in the presence of unknown levels of sample overlap. Univariate TWAS estimates are subsequently used as input to expand both of these matrices.

To create the *S*_*Full*_ matrix for multivariate TWAS within T-SEM, the *S*_*LDSC*_ matrix is expanded to include the (cis-) genetic covariance between the inferred gene expression and phenotypes, g_1_ through g_k_, by appending the vector *S*_*Gene*_:

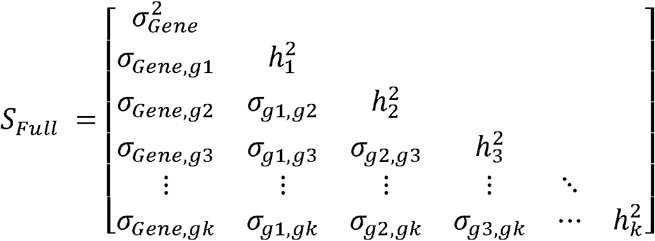

The 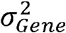 in the first cell of the matrix above is the (cis-) heritability of the expression of an individual gene, provided directly by FUSION.

The sampling covariance matrix, *V*_*SFull*_, associated with the expanded *S*_*Full*_ covariance matrix, consist of three blocks. The first block is the *V*_*SLDSC*_ matrix obtained from multivariable LDSC outlined above. The second block, *V*_*SGene*_, is composed of the sampling covariance matrix of the gene effects on the phenotypes. The sampling covariances of the gene-genotype covariances with one another are indexed using cross-trait LDSC intercepts. As these cross-trait intercepts reflect sampling correlations (weighted by sample overlap), they are rescaled relative to the sampling variances of the corresponding gene-genotype covariances to be on the scale of sampling covariances. We treat the sampling variance of the 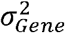 as fixed. In practice we set it to a very small value (1e-4), and set the sampling covariance between the 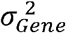 and all other terms to 0. The third, and final, block of the *V*_*SFull*_ matrix reflects the sampling covariance of the gene-genotype covariances from FUSION with the genetic variances and genetic covariances from multivariable LDSC. These sampling covariances are fixed to 0 as the gene-phenotype covariance will be independent of the individual SNP effects in all LD blocks except those which SNPs defining the gene occupy. As the sampling variance of the elements of *S*_*LDSC*_ reflect sampling variability with SNP test statistics in all LD blocks, their sampling covariance with the effect of a single gene is expected to approach 0. Taking these three components together, the *V*_*SFull*_ matrix for multivariate TWAS can be written in compact form as:

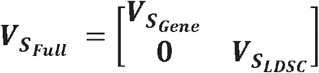

The *S*_*Full*_ and *V*_*SFull*_ matrices are constructed as many times as there are shared gene IDs across univariate FUSION outputs for individual traits.

### Scaling TWAS Output for T-SEM

The output from univariate summary-based TWAS (as estimated with FUSION^7^) is a predicted gene-trait Z-statistic that requires further transformation to be added to the *S*_*Full*_ matrix above. First, this Z-statistic is converted to a partially standardized regression coefficient and its *SE* using the formulas, 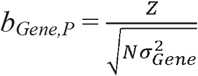 and 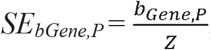, where 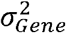 reflects the heritability estimate for an individual gene provided by FUSION. We refer to these as partially standardized regression coefficients as they are standardized relative to the variance in the outcome (i.e., the phenotype of interest), but not the predictor (i.e., a given gene). These partially standardized coefficients are subsequently transformed into gene-phenotype covariances (*σ* _*Gene,gk*_), as in the the *S*_*Full*_ matrix above, by multiplying by the variance (heritability) of the gene 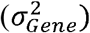.

### Q_Gene_ Test of Heterogeneity

Q_Gene_ indexes whether a given gene is more likely to operate through an identified common factor or via the independent pathways of the individual traits that define the factor. This metric helps to guard against identifying genes as operating through the common factor when they are, in fact, highly specific to a trait or subset of traits. That is, it formally tests the null hypothesis that the gene acts through a given factor. Q_Gene_ is calculated here using a two-step procedure that mirrors the steps outlined for Q_SNP_.^3,4^ In Step 1, a *common pathway* model is fit where the gene effect on the common factor, the (residual) variances of the factor and its indicators, and all but one factor loading that is scaled to unity for identification, are freely estimated. In Step 2, a *common plus independent pathways model* is fit where the factor loadings and the gene effect on the common factor are fixed from the parameter estimates in Step 1, and the direct effects of the gene on the indicators and the residual variances are freely estimated. Supplementary Figure 1b depicts this two-step procedure, as applied to the *g*-factor model, with parameters that are fixed in Step 2 depicted in red and those that are freely estimated in Step 2 depicted in black. The same formula used for estimates of model χ^2^ in Genomic SEM are used here to produce a χ^2^ distributed Q_Gene_ test statistic with degrees of freedom (*df*) equal to *k* −1, where *k* reflects the number of included phenotypes. For comparative purposes throughout the paper, we generally scale this Q_Gene_ test statistic to be a χ^2^ statistic with *df* = 1.

### T-SEM Simulations

#### Simulation Procedure

As a first step in the simulation procedure, the model implied genetic covariance matrix was calculated for the model in which the top gene expression hit from the real data analyses, ZSCAN9 in the cerebellum tissue, was specified to predict the *g-*factor. This reflects a best-case scenario for identifying common factor hits in T-SEM in that the model implied matrix reflects a pattern of relationships that operate entirely through the common factor. Seven versions of this model implied matrix were subsequently used to construct population generating covariance matrices, from which 250 covariance matrices were then sampled using *rmvnorm* in R for each of the seven scenarios (i.e., 1,750 total simulations). These seven scenarios consisted of: Scenario 1, reflective of the alluded to best-case scenario where the model implied matrix was unchanged; Scenario 2 in which the covariance between the gene and RT (the indicator with the smallest factor loading) was set to 0; Scenario 3 in which the covariance between the gene and Trails-b (the indicator with the largest factor loading) was set to 0; Scenario 4 in which the covariance between the gene and all indicators *except* trails-B was set to 0; Scenario 5 in which the covariance between the gene and all indicators *except* RT was set to 0; Scenario 6 in which the covariance between the gene and all indicators was set to 0; and Scenario 7 in which the directionality of the covariance between the gene and matrices, memory, and RT was reversed.

The observed sampling covariance matrix, 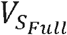, was used both to sample from the population matrices for each condition and was paired with each simulated genetic covariance matrix for the real data analyses. This has the advantage of providing a set of simulations that are both directly relevant to interpretation of the current analyses and, by definition, reflective of the types of real-data scenarios where T-SEM might be applied. All simulations applied the same Bonferroni corrected threshold for significance as used in the real-data analysis.

#### Simulation Results

These scenarios were selected to reflect a gradient of deviations from the factor model, where Scenario 1 exactly matches the common factor model and Scenario 7 reflects a strong departure from the model wherein the gene has directionally opposing effects across the *g*-factor indicators. This allowed us to test whether T-SEM appropriately down weights estimates of gene effects on the common factor in a graded fashion as the generating population gradually shifts further away from the common factor structure. Results for Scenario 1, with population gene effects specified to operate solely through the common factor, revealed hits on the common factor for all but 1 run (i.e., 99.6% hits; Supplementary Table 2) and evinced the strongest signal across scenarios (Supplementary Figure 4). The signal was slightly reduced for Scenario 2, in which the gene effect was set to 0 for the indicator with the smallest loading (RT) and more attenuated for Scenario 3, in which the gene effect was set to 0 for the indicator with the largest loading (Trails-b; Supplementary Figure 2). It is also worth noting that Scenario 3 still produced hits for 82.7% of the simulations. As expected, the signal was greatly reduced relative to these three scenarios for Scenario 4, in which the gene effect was set to 0 for all but Trails-b, and further reduced for Scenario 5, in which the gene effect was set to 0 for all but RT, with no simulations producing hits for either scenario. Taken together, this demonstrates that indicators with larger factor loadings are appropriately weighted when estimating gene effects *and* that the signal for the common factor is by no means a recapitulation of the signal for the strongest indicator. For Scenario 6, in which the gene effect in the population was 0 across all indicators, simulation results revealed a null signal. Finally, for Scenario 7, in which the directionality of the gene effect was reversed for three of the indicators, 35.1% of the runs were identified as significant for the common factor. However, all of these significant runs were also identified as hits for Q_Gene_, which we consider next.

The pattern of results of Q_Gene_ were also consistent with expectation. For Scenario 1, there was a null signal, with no simulations identified as Q_Gene_ hits (i.e., no false positives) consistent with the fact that the population did not deviate from the factor structure (Supplementary Figures 3 and 5). The next closest signal was for Scenario 6 in which all associations were set to 0 in the population, reflecting the fact that a pattern of null associations across the indicators largely aligns with a common factor model for highly correlated traits. Scenarios 2 and 5, in which the population effect of the gene on RT and all indicators except RT was set to 0, respectively, evinced a similar Q_Gene_ signal that was slightly stronger than Scenario 6, but did not produce any Q_Gene_ hits, reflecting the relatively attenuated influence of RT (the indicator with the smallest loading). Scenarios 3, in which the population effect of the gene on Trails-B was 0, evinced the next highest signal, with 39.4% of runs producing Q_Gene_ hits, followed by Scenario 4, in which the gene effect on all indicators except Trails-B was set to 0 in the population, with 46.1% of runs producing Q_Gene_ hits. As expected, Scenario 7, which deviated the strongest from the model with directionally opposing gene effects on matrices, memory, and RT relative to the remaining indicators, showed the strongest Q_Gene_ signal by far, with 99.6% of simulations identifying a Q_Gene_ hit.

### Quality Control Procedures

We refer the reader to the original article describing genetic *g* for details about sample ascertainment, quality control, and related procedures for the seven cognitive tests,^4^ in addition to the corresponding articles for schizophrenia^21^ and bipolar disorder.^22^ Default quality control (QC) procedures were used for the *munge* function in Genomic SEM prior to running either LDSC or S-LDSC. This included removing SNPs with an MAF < 1%, information scores (INFO) < .9, SNPs from the MHC region, and filtering SNPs to HapMap3. The LD scores used for the overall LDSC model were estimated from the European sample of 1000 Genomes. Details on the LD scores stratified by functional annotations used for S-LDSC are provided in further detail below. LD scores used as input for both LDSC and S-LDSC were also restricted to HapMap3 SNPs as these tend to be well-imputed and produce accurate estimates of heritability.

Prior to running FUSION, alleles were aligned across univariate summary statistics to the 1000 Genomes Phase 3 LD reference panel, restricted to SNPS with an MAF > 1%, SNPs with an INFO score < 0.6, and restricted to those SNPs that were present across all seven cognitive tests. Using these QC steps, there were 7,857,346 SNPS present across all tests, of which 1,157,709 SNPs were present in the LD reference panel data and subsequently used by FUSION to produce univariate TWAS estimates. Univariate FUSION summary statistics were subsequently standardized with respect to the total variance in the outcome using the *read_fusion* function in *GenomicSEM*. Standard errors were also corrected for genomic inflation using the conservative approach of multiplying the standard errors by the univariate LDSC intercept when the intercept was above 1.

### Univariate TWAS

The FUSION software^7^ was used to perform transcriptomic imputation (TI)/summary-based univariate TWAS for the seven cognitive summary statistics. Functional weights were used from eQTL reference panels for 13 brain tissue panels from the Genotype-Tissue expression project v7 (GTEx; n = 753; https://gtexportal.org/home/datasets)^5^ and 2 dlPFC panels from the CommonMind Consortium (CMC; n = 452; Supplementary Table 7).^6^ The functional weights utilized in the current study are all publicly available on the FUSION website and were pre-computed using the package defaults. These weights were coupled with LD information from the 1000 Genomes v3 European subsample to produce univariate TWAS test statistics. The FUSION package quality control defaults were also used for summary-based TWAS, including a minimum R^2^ imputation accuracy of .7 per gene and a maximum of 50% of SNPs allowed to be missing per gene. Using these defaults, results were not calculated for 233 genes, for a remainder of 52,849 genes across the 15 tissues. A strict Bonferroni-corrected threshold was used for both the *g*-factor T-SEM results and Q_Gene_ test statistics using an FDR of .05 (.05/52,849).

TWAS test statistics produced by FUSION are well-calibrated under the null of no GWAS association, but can become inflated as a result of random quantitative trait loci (QTL) colocalization. This can occur when a locus is both highly significant and characterized by extensive LD. To guard against these instances, FUSION offers a permutation test statistic that recomputes the TWAS test statistic conditional on the GWAS effects at that locus after randomly reordering the QTL weights. This permutation test asks whether the distribution of QTL effect sizes is *by itself* sufficient for producing a significant TWAS association. The output is an empirically derived *p*-value that indexes the proportion of permutations that were more significant than the observed TWAS *p*-value. For the current analyses, 100,000 permutations were run per gene for all genes. These univariate, empirical *p*-values were then used as input for a separate, multivariate TWAS of the *g*-factor in Genomic SEM. It is important to note that these univariate empirical *p*-values, and consequently the multivariate TWAS results, are highly conservative such that genes that are truly causal in the population may fail to reject the null when their QTLs are characterized by extensive and complex patterns of LD. At the same time, genes that remain significant for the permutation test can be interpreted as less likely to be colocalized due to chance.

### T-SEM Follow-up Analyses

All follow-up analyses were computed at the level of the *g*-factor, as opposed to at the level of the cognitive indicators used as input for a separate multivariate TWAS. This is due to the fact that these approaches are recommended and coded to be used for significant genes only. That is, by restricting to listwise deleted significant genes across the indicators, the end result would be an overly attenuated set of genes that would not be appropriate as input for separate T-SEM analyses.

#### Conditional Analyses

As many genes are present across tissue types, and genes overlap in physical proximity, it can also be useful to consider the conditional effect of each gene. Conditional analyses are conducted in FUSION via an iterative procedure that adds predictors to the model until no significant associations remain. Analyses were conducted using the package default locus window of 100,000 bp. It is of note that genes that are estimated to be jointly significant are not necessarily more likely to be causal than those that are not jointly significant. This is due to the fact that the gene expression features in the latter case may simply be characterized by high correlations with other features. In addition, it is useful to consider to what extent the model explains observed SNP effects by examining SNP effects conditional on TWAS estimates. In this context, it is informative to examine both the level of significance of the top SNP within a region before and after conditioning on TWAS estimates along with the proportion of variance explained in that region by corresponding TWAS results. If TWAS estimates explain a small proportion of the GWAS variance in a region, this suggests that TWAS estimates are tagging an independent causal feature, with the inverse being true when large amounts of variance are explained.

#### Colocalization Analyses

Colocalization analyses can also be used to examine the probability of a shared causal variant between gene expression and the trait of interest (i.e., whether there are colocalized functional and GWAS associations). This reflects an alternative to a TWAS, which examines the evidence of a signification association between imputed gene expression and the trait. Bayesian colocalization analyses were conducted using the *coloc* R package^23^ run through FUSION. When implemented via FUSION, the *coloc* package works by estimating the posterior probability of different configurations of a single causal SNP for both gene expression and the trait, along with the posterior probability that the gene expression and trait share these configurations. The output is posterior probabilities for five scenarios. Model 0 (PP0 in Table S2) reflects a situation in which there is no GWAS or functional association. Model 1 (PP1) examines the probability of a functional association only and Model 2 (PP2) examines the probability of a GWAS association only. Model 3 (PP3) examines whether there are independent functional and GWAS associations. Finally, Model 4 (PP4) examines the probability of colocalized functional and GWAS associations. As the *coloc* software assumes a single causal variant, and FUSION models assume multiple eQTLs, a low posterior probability of Model 3, as opposed to a high posterior probability of Model 4, can also be taken as good indication of colocalization. These posterior probabilities were calculated using the *g*-factor GWAS summary statistics and functional reference weights across tissues as input.

#### Gene-set analyses

GeneNetwork v2.0^24^ was used to estimate gene co-expression networks in order to better characterize the multivariate TWAS results for both the *g*-factor and Q_Gene._ Genes used as input for the *g*-factor to create the co-expression network included those genes that were significant at a Bonferroni corrected threshold for 52,849 tests and did not overlap with significant Q_Gene_ hits for the same gene and tissue type. These genes were restricted still further to those unique gene IDs across tissue types for a total of 76 genes used as input. There were 62 unique genes for Q_Gene,_ among which 3 were not present in the database, for a total of 59 gene IDs used as input. Pathways were subsequently analyzed across 3,033 pathways from the Reactome database and the three primary Gene Ontology (GO) databases for biological processes, molecular functions, and cellular components. A Bonferroni corrected threshold was used to identify significant pathways for 3,033 tests at an FDR of .05 (i.e., p < 1.65e-5).

### Stratified Genomic SEM

Stratified Genomic SEM begins by estimating genetic correlation and covariance matrices stratified across different gene sets and categories (referred to as functional annotations) using a multivariable version of Stratified LDSC.^14,18^ The model of interest is then estimated for the functional annotation including all SNPs. In the context of the current analyses—where the parameters of interest reflect the factor variance of *g*, genetic overlap across *g* and the psychotic disorders factor, and the residual variances of the indicators—the factor loadings are subsequently fixed from the estimates obtained from the annotation including all SNPs, and the remaining model parameters are freely estimated within each annotation. The freely estimated parameter and sandwich corrected standard error within a given annotation are then scaled relative to the estimate obtained for the annotation including all SNPs, such that the estimate now reflects a proportion of the total, genome-wide estimate. In the unstandardized case, the enrichment “ratio of ratios” is then calculated by scaling this proportional estimate by the number of SNPs in the corresponding annotation as a proportion of the total SNPs examined. Enrichment is then observed when the proportion of genome-wide variance observed in an annotation is greater than the proportional size of the annotation. In the standardized case (e.g., for the current analyses when stratified genetic correlation matrices are used as input or factor correlations are examined) the estimate is not rescaled by the proportional size of the annotation as all annotations, including the genome-wide annotation, are on the same scale.

Zero-order, stratified genetic covariance and correlation matrices were estimated using the *s_ldsc* function in Genomic SEM. This included 97 functional annotations from the 1000 Genomes Phase 3 BaselineLD Version 2.2 provided by the original S-LDSC authors,^18^ tissue specific histone marks from the Roadmap Epigenetics Project,^25^ and tissue specific gene expression from GTEx^5^ and DEPICT.^26^ For tissue specific gene expression and histone/chromatin marks we utilized only brain and endocrine relevant regions in addition to 5 randomly selected control regions from each (i.e., 10 controls total). We additionally utilized 29 functional annotations created using data from Genome Aggregation Database (gnomAD)^27^ and GTEx^28^ to examine the interaction between protein-truncating variant (PTV)-intolerant (PI) genes and human hippocampal and prefrontal brain cells. Details on parameters used to create these 29 annotations can be found in Grotzinger et al., 2020.^14^

Enrichment was not estimated for continuous or flanking window/control annotations, yielding a total of 168 binary annotations. We further remove 13 annotations that were non-positive definite and required smoothing the stratified covariance matrix such that any point estimate in the matrix produced a Z-statistic discrepancy > 1.96 pre- and post-smoothing. For a Bonferroni correction at < .05 this corresponds to *p* < 3.22E-4 across the 155 remaining functional annotations. Analyses examining enrichment of the genetic correlation between a *g*-factor with a psychotic disorder factor largely mirrored those for enrichment of the *g*-factor. When pruning based on the Z-statistic discrepancy for smoothing, a total of 15 annotations were removed for this analysis; however, we use the same Bonferroni corrected threshold for 155 tests for comparative purposes.

## Data Availability

The data that support the findings of this study are all publicly available or can be requested for access. Specific download links for various datasets are directly below.
Summary statistics for the g-factor and the seven, individual cognitive traits are available from:
https://datashare.is.ed.ac.uk/handle/10283/3756
Summary statistics for data from the Psychiatric Genomics Consortium (PGC) can be downloaded or requested here:
https://www.med.unc.edu/pgc/download-results/
Data from gnomAD used to identify PI genes for creation of annotations can be downloaded here: https://storage.googleapis.com/gnomad-public/release/2.1.1/constraint/gnomad.v2.1.1.lof_metrics.by_gene.txt.bgz
Gene count data per cell for creation of annotations were obtained from: https://storage.googleapis.com/gtex_additional_datasets/single_cell_data/GTEx_droncseq_hip_pcf.tar
Data which maps individual cells to cell types (e.g. neuron, astrocyte etc.) were obtained from: https://static-content.springer.com/esm/art%3A10.1038%2Fnmeth.4407/MediaObjects/41592_2017_BFnmeth4407_MOESM10_ESM.xlsx
Links to the LD-scores, reference panel data, and the code used to produce the current results can all be found at: https://github.com/GenomicSEM/GenomicSEM/wiki
Links to the BaselineLD v2.2 annotations can be found here:
https://data.broadinstitute.org/alkesgroup/LDSCORE/
Links to the reference weights used for FUSION from GTEx and CMC can be found here:
http://gusevlab.org/projects/fusion/

http://gusevlab.org/projects/fusion/

https://github.com/GenomicSEM/GenomicSEM/wiki

https://static-content.springer.com/esm/art%3A10.1038%2Fnmeth.4407/MediaObjects/41592_2017_BFnmeth4407_MOESM10_ESM.xlsx

https://storage.googleapis.com/gtex_additional_datasets/single_cell_data/GTEx_droncseq_hip_pcf.tar

https://storage.googleapis.com/gnomad-public/release/2.1.1/constraint/gnomad.v2.1.1.lof_metrics.by_gene.txt.bgz

https://www.med.unc.edu/pgc/download-results/

https://datashare.is.ed.ac.uk/handle/10283/3756

## Code Availability

GenomicSEM software (which now includes the T-SEM and Stratified GenomicSEM extensions), is an R package that is available from GitHub at the following URL: https://github.com/GenomicSEM/GenomicSEM

Directions for installing the GenomicSEM R package can be found at: https://github.com/GenomicSEM/GenomicSEM/wiki

## Data Availability

The data that support the findings of this study are all publicly available or can be requested for access. Specific download links for various datasets are directly below.

Summary statistics for the *g-*factor and the seven, individual cognitive traits are available from: https://datashare.is.ed.ac.uk/handle/10283/3756

Summary statistics for data from the Psychiatric Genomics Consortium (PGC) can be downloaded or requested here:

https://www.med.unc.edu/pgc/download-results/

Data from gnomAD used to identify PI genes for creation of annotations can be downloaded here: https://storage.googleapis.com/gnomad-public/release/2.1.1/constraint/gnomad.v2.1.1.lof_metrics.by_gene.txt.bgz

Gene count data per cell for creation of annotations were obtained from: https://storage.googleapis.com/gtex_additional_datasets/single_cell_data/GTEx_droncseq_hip_pcf.tar

Data which maps individual cells to cell types (e.g. neuron, astrocyte etc.) were obtained from: https://static-content.springer.com/esm/art%3A10.1038%2Fnmeth.4407/MediaObjects/41592_2017_BFnmeth4407_MOESM10_ESM.xlsx

Links to the LD-scores, reference panel data, and the code used to produce the current results can all be found at: https://github.com/GenomicSEM/GenomicSEM/wiki

Links to the BaselineLD v2.2 annotations can be found here: https://data.broadinstitute.org/alkesgroup/LDSCORE/

Links to the reference weights used for FUSION from GTEx and CMC can be found here: http://gusevlab.org/projects/fusion/

## Acknowledgements

This work presented here would not have been possible without the enormous efforts put forth by the investigators and participants from Psychiatric Genetics Consortium and UK Biobank. The work from these contributing groups was supported by numerous grants from governmental and charitable bodies as well as philanthropic donation. Research reported in this publication was supported by the National Institute Of Mental Health of the National Institutes of Health under Award Number R01MH120219. The content is solely the responsibility of the authors and does not necessarily represent the official views of the National Institutes of Health. ADG was additionally supported by NIH Grant R01HD083613. EMTD was additionally supported by NIH grants R01AG054628 and R01HD083613 and the Jacobs Foundation. EMTD is a faculty associate of the Population Research Center at the University of Texas, which is supported by NIH grant P2CHD042849. Additionally, EMTD is a member of the and Center on Aging and Population Sciences (CAPS) at the University of Texas at Austin, which is supported by NIH grant P30AG066614. MGN is additionally supported by ZonMW grants 849200011 and 531003014 from The Netherlands Organization for Health Research and Development, a VENI grant awarded by NWO (VI.Veni.191G.030) and is a Jacobs Foundation Fellow.

