## Supplementary material for "Transcriptome-wide and Stratified Genomic Structural Equation Modeling Identify Neurobiological Pathways Underlying General and Specific Cognitive Functions": Online Supplement

### Supplementary Method

#### Enrichment in Standardized Space

We went on to estimate enrichment for *g* using stratified genetic correlation (as opposed to covariance) matrices, which can be used to identify annotations for which enrichment of pleiotropic and trait-specific signal are disproportionate. As we highlight in the **Online Method**, estimation of enrichment in standardized space (e.g., when using stratified correlation matrices as input) does not require diving the enrichment estimate by the proportional size of the annotation, as all annotations (including the genome-wide annotation) are on the same scale. We did not observe any significantly enriched annotations for these stratified correlation analyses (Supplementary Table 8; Supplementary Figure 14). We also did not observe any significant enrichment of the genetic correlation between *g* and the psychotic disorders factors (Supplementary Table 8). We note that these analyses are likely underpowered as significant enrichment in a standardized space requires an annotation that indexes genetic overlap across traits far above and beyond what is observed at the genome-wide level. Therefore, we would characterize our results as reflecting clear genetic risk-sharing within a number of annotations, but inconclusive as to whether these annotations are relevant to both pleiotropic and trait-specific signal.

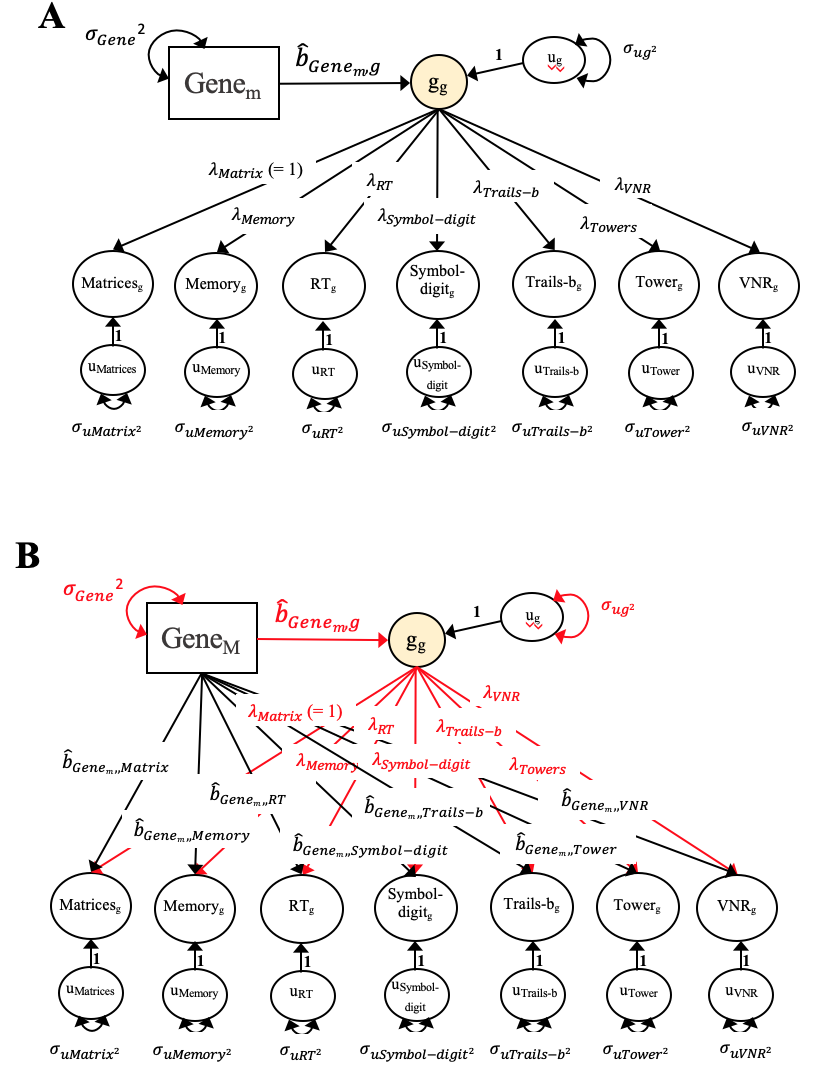

Figure S1. T-SEM Models. *Panel A* depicts the model run to obtain estimates of each gene on the g-factor. This model is run *M* times to reflect the number of genes present across all traits, in this case 52,849 genes. *Panel B* depicts the model used to estimate Q_Gene_, where Red lines and parameters are fixed from Step 1, and black lines and parameters are freely estimated in Step 2. The loading of the first indicator for each factor is fixed to 1 in all panels for identification purposes.

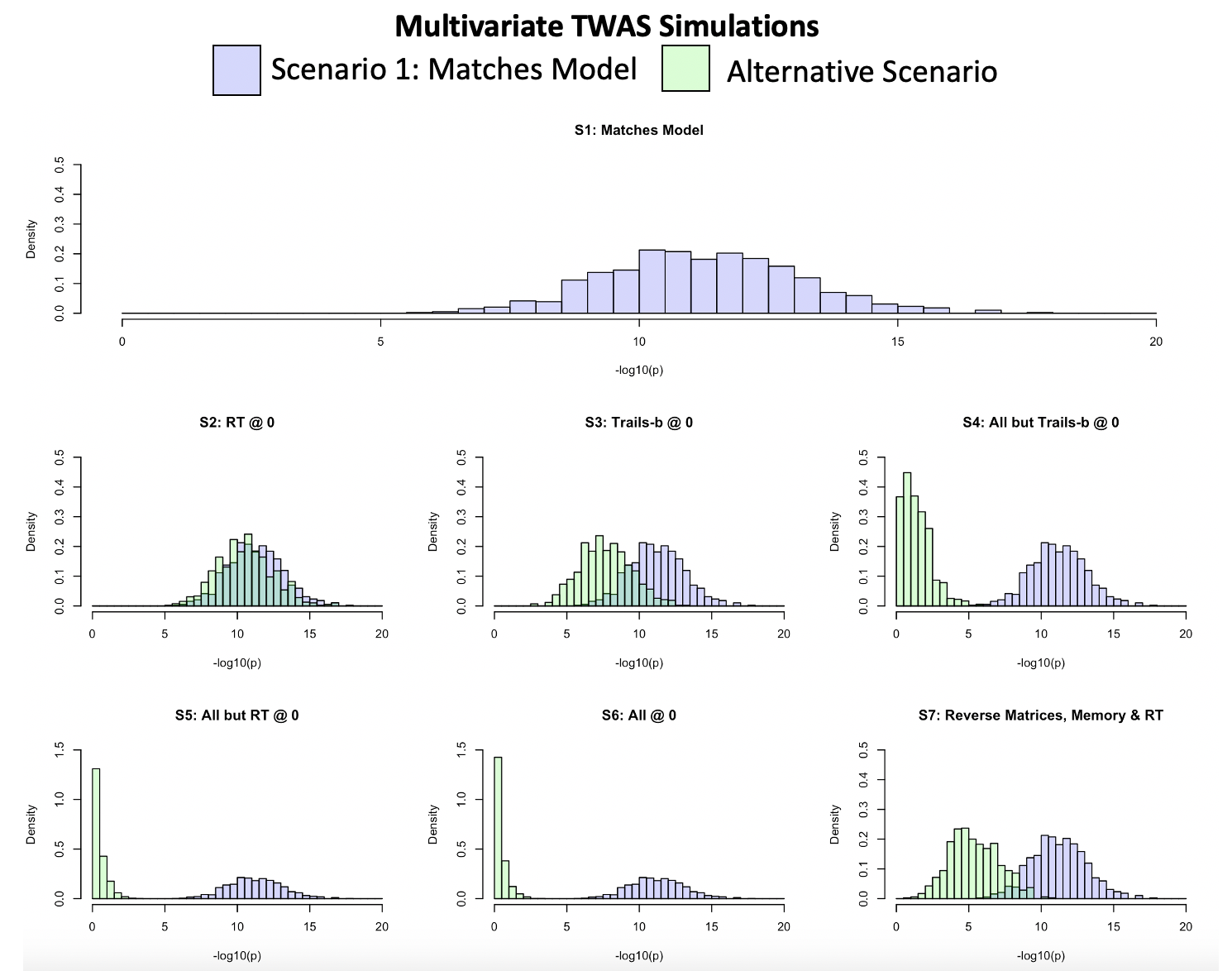

Figure S2. Histograms of T-SEM Simulation Results. Panels depict the -log10(p) values for estimated gene effects on the g-factor across the 7 different population generating scenarios. All panels depict in blue as a reference point the simulation scenario that exactly matched the factor model (i.e., Scenario 1 depicted in upper panel) and in green the specific scenario indicated in the histogram title.

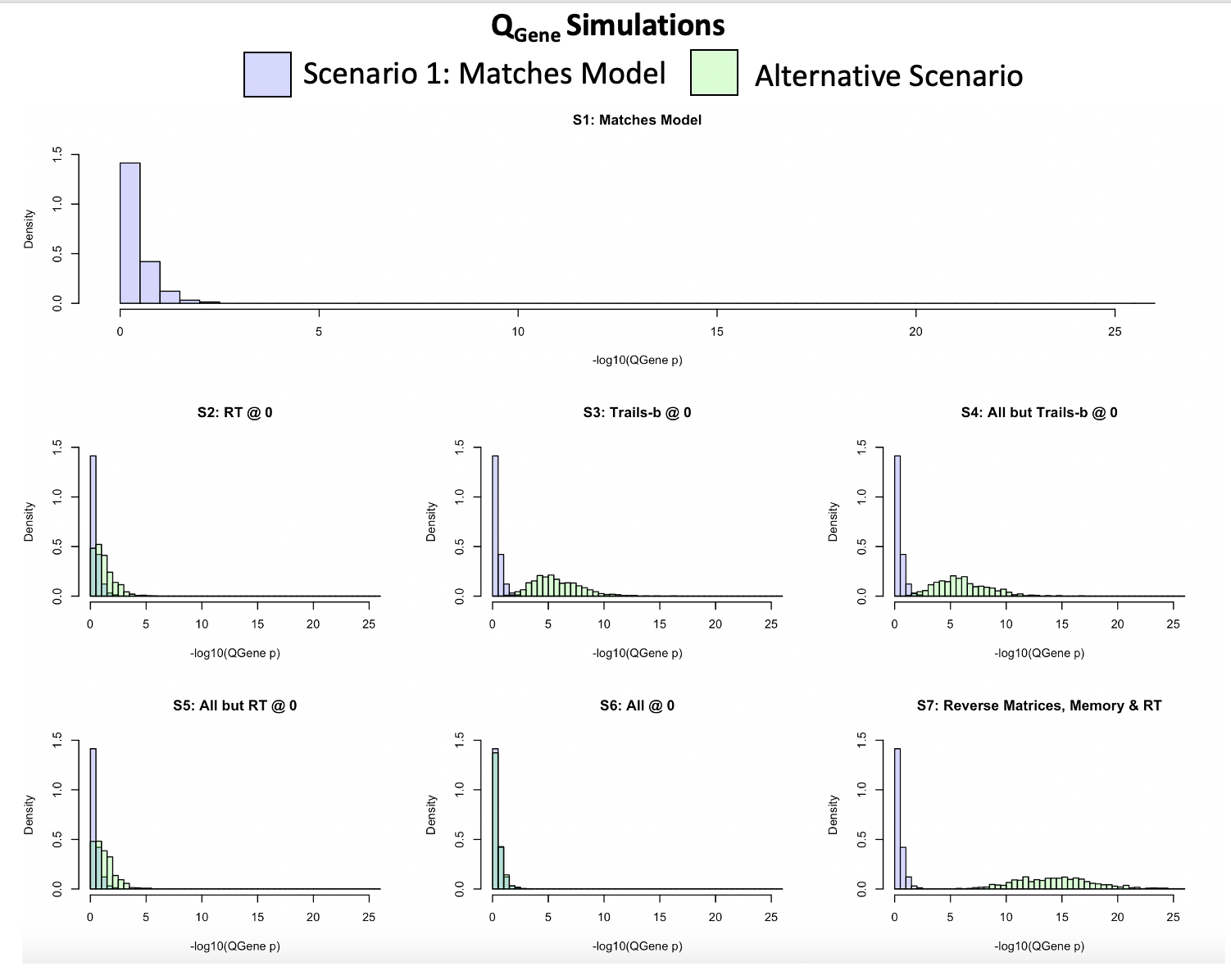

Figure S3. Histograms of Q_Gene_ Simulation Results. Panels depict the -log10(p) values for Q_Gene_ across the 7 different population generating scenarios. All panels depict in blue as a reference point the simulation scenario that exactly matched the factor model (i.e., Scenario 1 depicted in upper panel) and in green the specific scenario indicated in the histogram title.

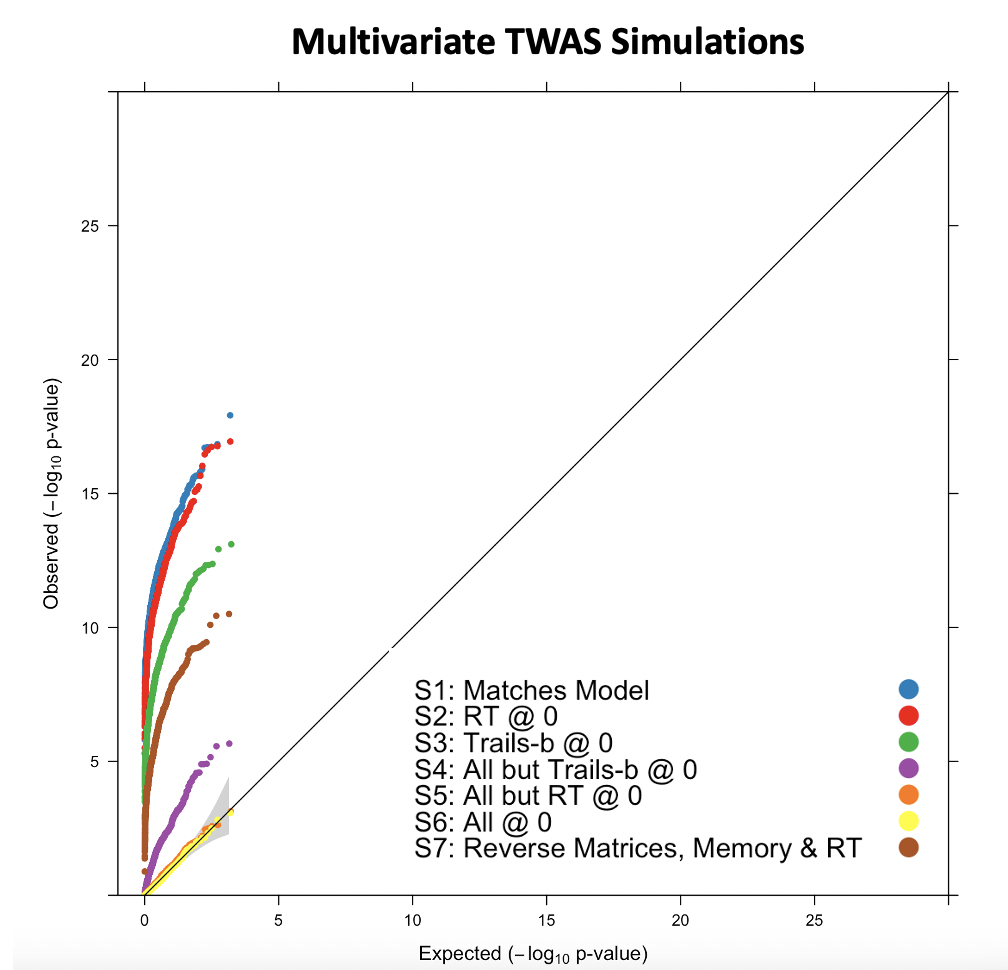

Figure S4. QQ-plot of T-SEM Simulation Results. QQ-plot depicts the -log10(p) values for gene effects on the g-factor for the 7 different population generating scenarios with: Scenario 1 that matches the factor model depicted in blue; Scenario 2 with the covariance between the gene and reaction time (RT) set at 0 in the generating population in red; Scenario 3 with the covariance between the gene and Trails-b set at 0 in the generating population in green; Scenario 4 with the covariance between the gene and all traits except Trails-b set at 0 in the in the generating population in purple; Scenario 5 with the covariance between the SNP and all traits except RT at 0 in the generating population in orange; Scenario 6 with the covariance between the gene and all cognitive traits set at 0 in the generating population in yellow; and Scenario 7 with the covariance between the gene and Matrices, Memory, and RT traits directionally reversed in brown.

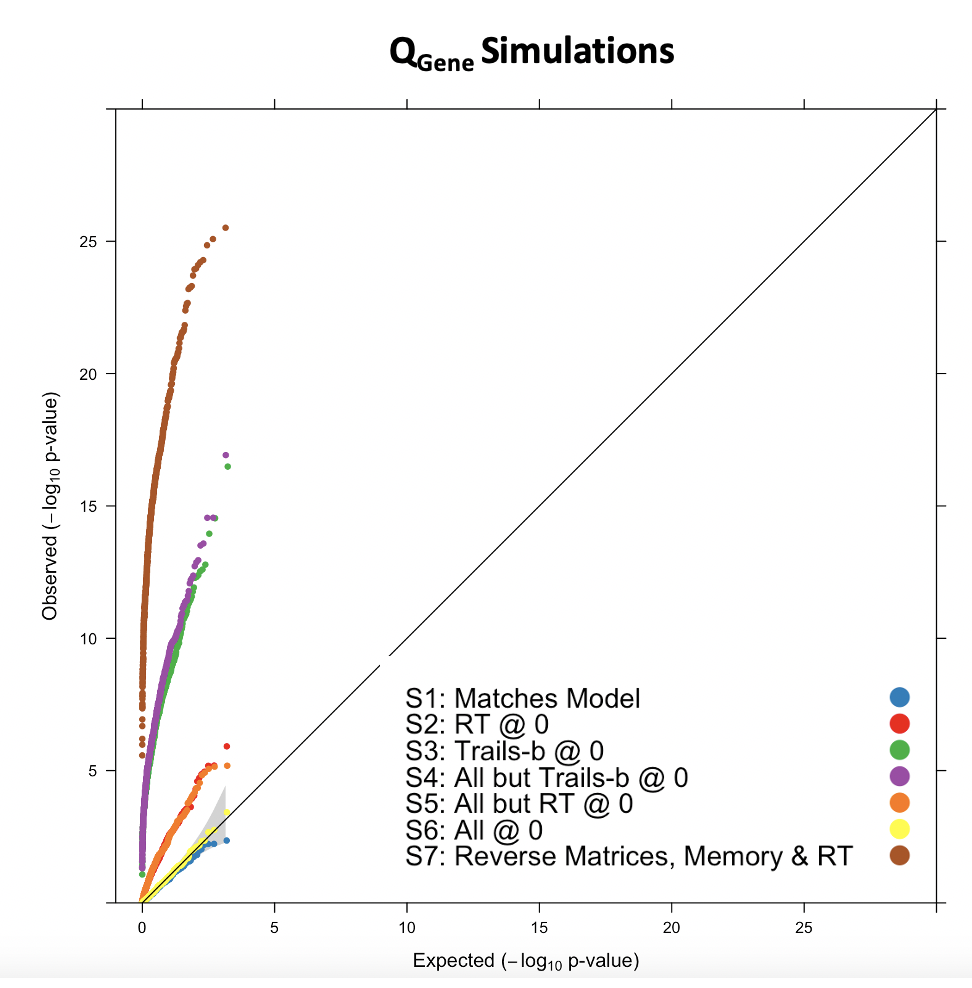

Figure S5. QQ-plot of Q_Gene_ Simulation Results. QQ-plot depicts the -log10(p) values for Q_Gene_ the 7 different population generating scenarios with: Scenario 1 that matches the factor model depicted in blue; Scenario 2 with the covariance between the gene and reaction time (RT) set at 0 in the generating population in red; Scenario 3 with the covariance between the gene and Trails-b set at 0 in the generating population in green; Scenario 4 with the covariance between the gene and all traits except Trails-b set at 0 in the in the generating population in purple; Scenario 5 with the covariance between the SNP and all traits except RT at 0 in the generating population in orange; Scenario 6 with the covariance between the gene and all cognitive traits set at 0 in the generating population in yellow; and Scenario 7 with the covariance between the gene and Matrices, Memory, and RT traits directionally reversed in brown.

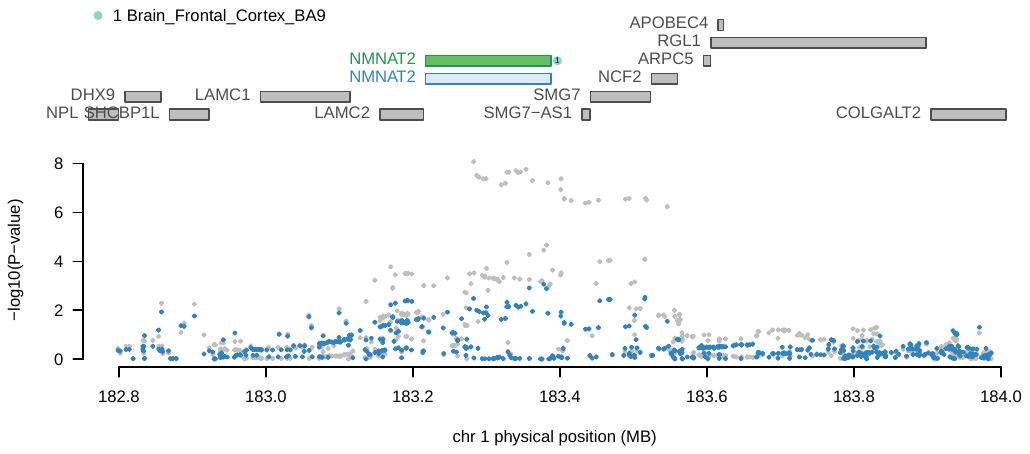

Figure S6a. Regional Association Plot for Locus 1 from g-factor Conditional Analyses. The top half of the panel displays all of the genes located in the locus window. Genes that were marginally significant are highlighted in blue while those that were jointly significant are highlighted in green. The color and numbering of the point next to jointly significant green genes indicates the specific tissue type (legend in upper-left) for which that gene was jointly significant. The bottom half depicts the Manhattan plot of the GWAS SNP effects within the locus prior to (grey) and after (blue) conditioning on the jointly significant genes in green. There were no genome-wide significant effects within this locus after conditioning on the predicted expression of NMNAT2 in the frontal cortex.

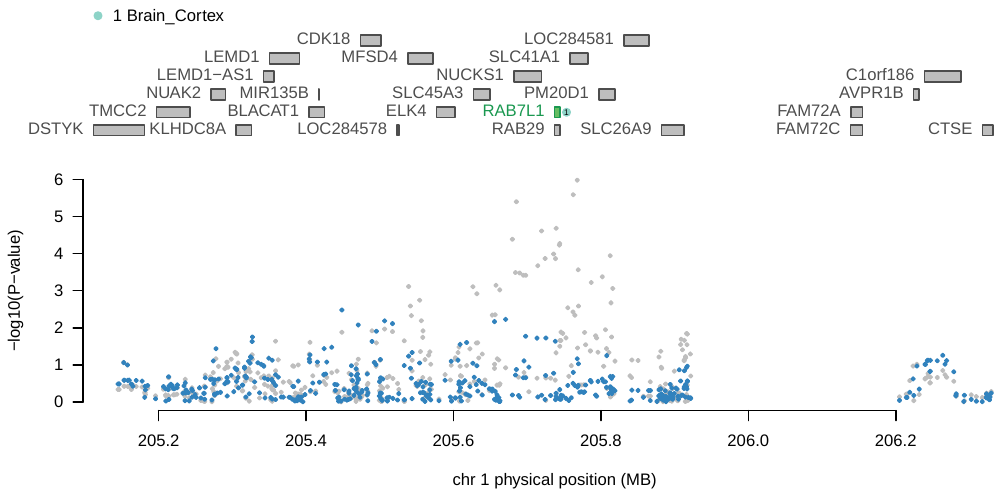

Figure S6b. Regional Association Plot for Locus 2 from g-factor Conditional Analyses. The top half of the panel displays all of the genes located in the locus window. Genes that were marginally significant are highlighted in blue while those that were jointly significant are highlighted in green. The color and numbering of the point next to jointly significant green genes indicates the specific tissue type (legend in upper-left) for which that gene was jointly significant. The bottom half depicts the Manhattan plot of the GWAS SNP effects within the locus prior to (grey) and after (blue) conditioning on the jointly significant genes in green. There were no marginally significant genome-wide effects within this locus after conditioning on the predicted expression of RAB7L1 in the cortex.

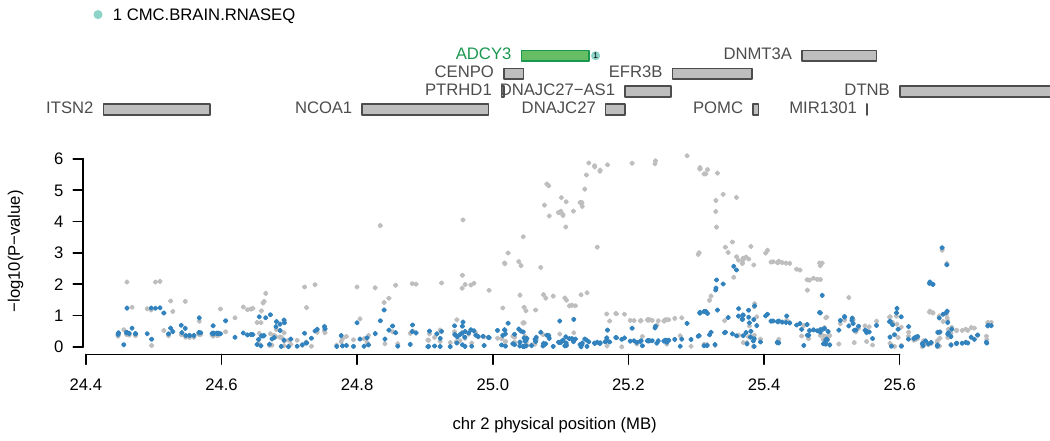

Figure S6c. Regional Association Plot for Locus 3 from g-factor Conditional Analyses. The top half of the panel displays all of the genes located in the locus window. Genes that were marginally significant are highlighted in blue while those that were jointly significant are highlighted in green. The color and numbering of the point next to jointly significant green genes indicates the specific tissue type (legend in upper-left) for which that gene was jointly significant. The bottom half depicts the Manhattan plot of the GWAS SNP effects within the locus prior to (grey) and after (blue) conditioning on the jointly significant genes in green. There were no marginally significant genome-wide effects within this locus after conditioning on the predicted expression of ADCY3 in the Common Mind Consortium RNA-seq dlPFC tissue type.

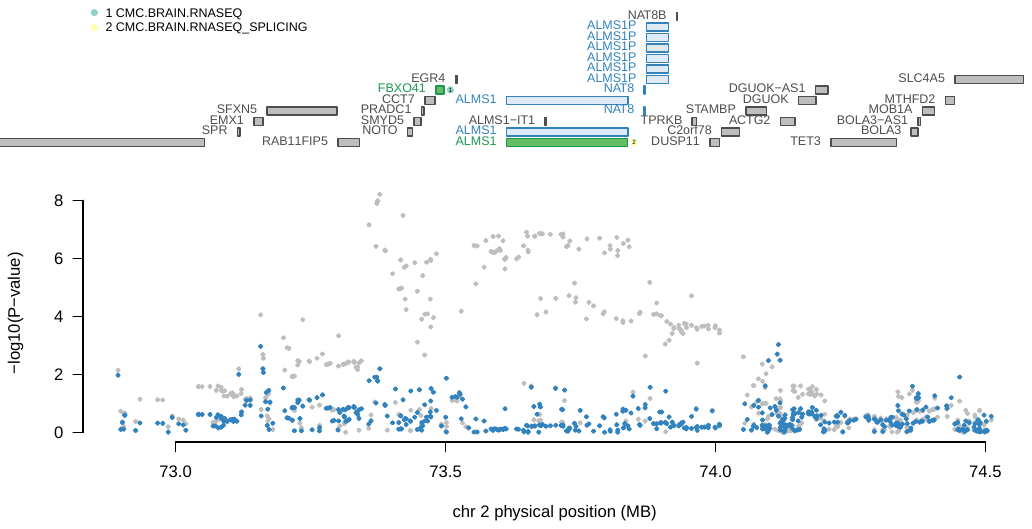

Figure S6d. Regional Association Plot for Locus 4 from g-factor Conditional Analyses. The top half of the panel displays all of the genes located in the locus window. Genes that were marginally significant are highlighted in blue while those that were jointly significant are highlighted in green. The color and numbering of the point next to jointly significant green genes indicates the specific tissue type (legend in upper-left) for which that gene was jointly significant. The bottom half depicts the Manhattan plot of the GWAS SNP effects within the locus prior to (grey) and after (blue) conditioning on the jointly significant genes in green. There were no significant genome-wide effects within this locus after conditioning on the predicted expression of FBXO41 gene in the Common Mind Consortium RNA-seq dlPFC tissue type and the ALMS1 gene in the Common Mind Consortium RNA-seq splicing dlPFC tissue type.

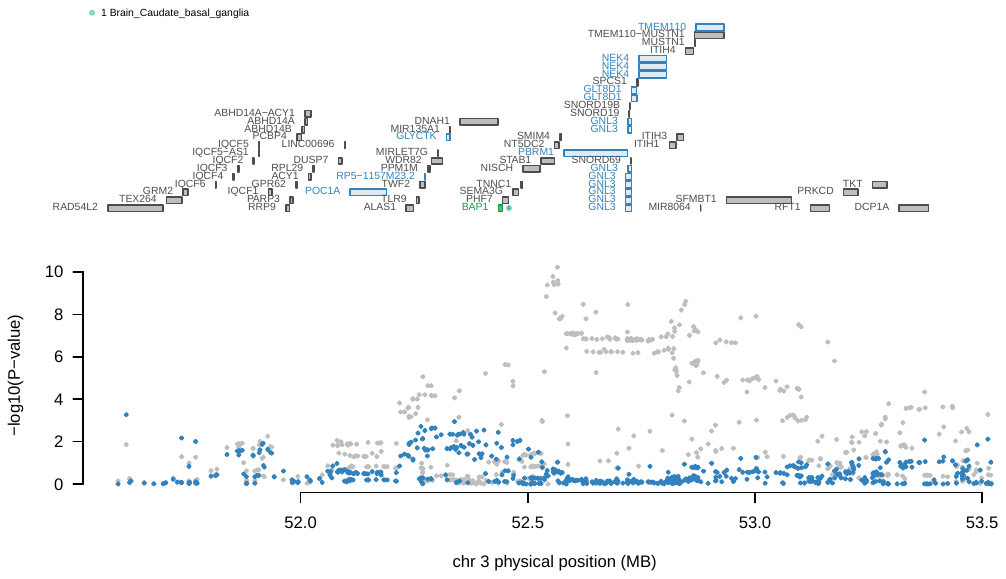

Figure S6e. Regional Association Plot for Locus 6 from g-factor Conditional Analyses. The top half of the panel displays all of the genes located in the locus window. Genes that were marginally significant are highlighted in blue while those that were jointly significant are highlighted in green. The color and numbering of the point next to jointly significant green genes indicates the specific tissue type (legend in upper-left) for which that gene was jointly significant. The bottom half depicts the Manhattan plot of the GWAS SNP effects within the locus prior to (grey) and after (blue) conditioning on the jointly significant genes in green. There were no significant genome-wide effects within this locus after conditioning on the predicted expression of BAP1 gene in the caudate tissue type.

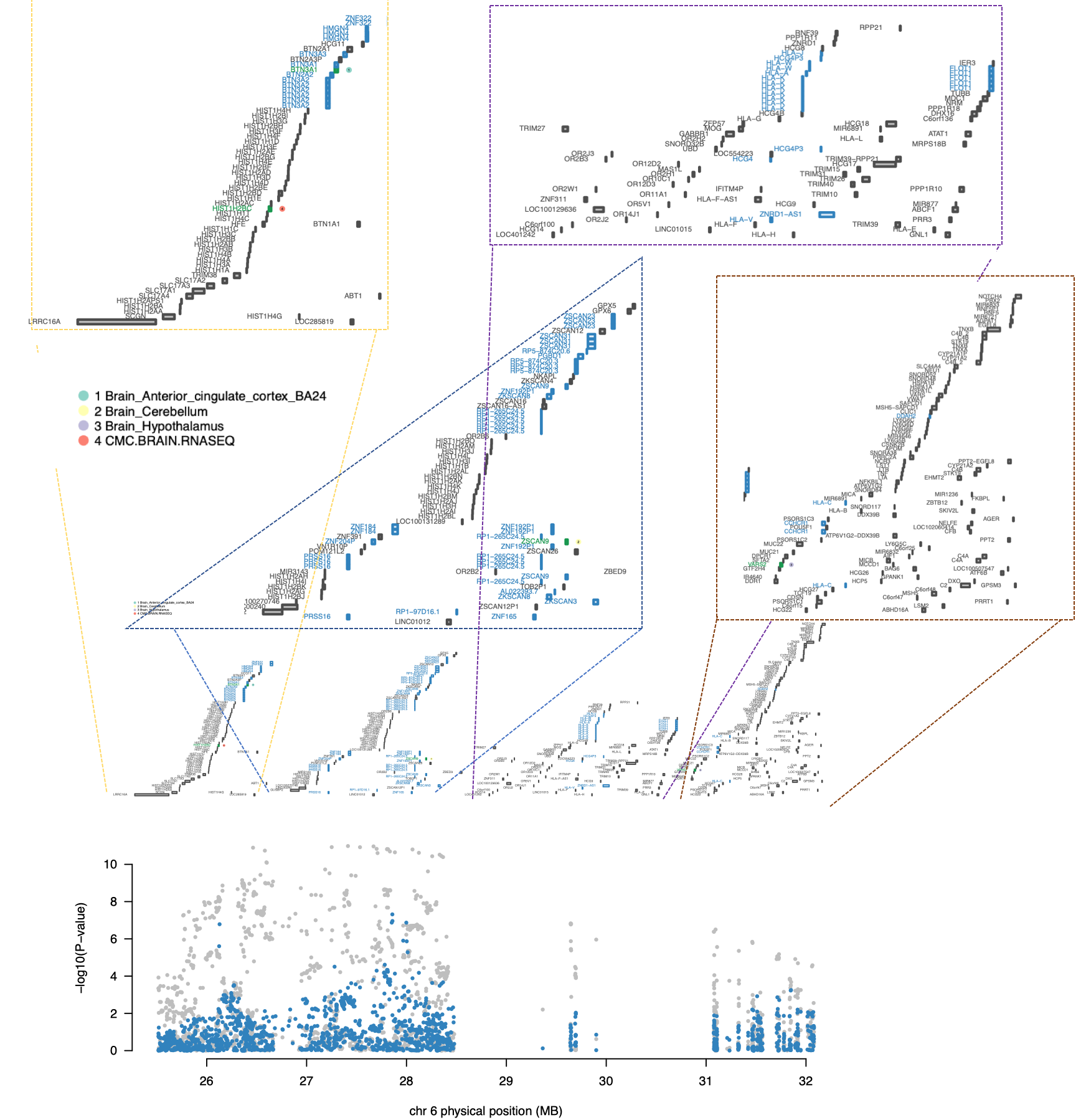

Figure S6f. Regional Association Plot for Locus 7 from g-factor Conditional Analyses. The top half of the panel displays all of the genes located in the locus window. Genes that were marginally significant are highlighted in blue while those that were jointly significant are highlighted in green. The color and numbering of the point next to jointly significant green genes indicates the specific tissue type (legend in upper-left) for which that gene was jointly significant. The bottom half depicts the Manhattan plot of the GWAS SNP effects within the locus prior to (grey) and after (blue) conditioning on the jointly significant genes in green. There were no significant genome-wide effects within this locus after conditioning on the predicted expression of HIST1H2Bc gene in the Common Mind Consortium RNA-seq dlPFC tissue type tissue type, the ZSCAN9 gene in the cerebellum, the VARS2 gene in the hypothalamus and the BTN3A1 gene in the anterior cingulate cortex. Due to the number of genes within this particular locus, zoomed in portions of the top half of the plot are provided directly above.

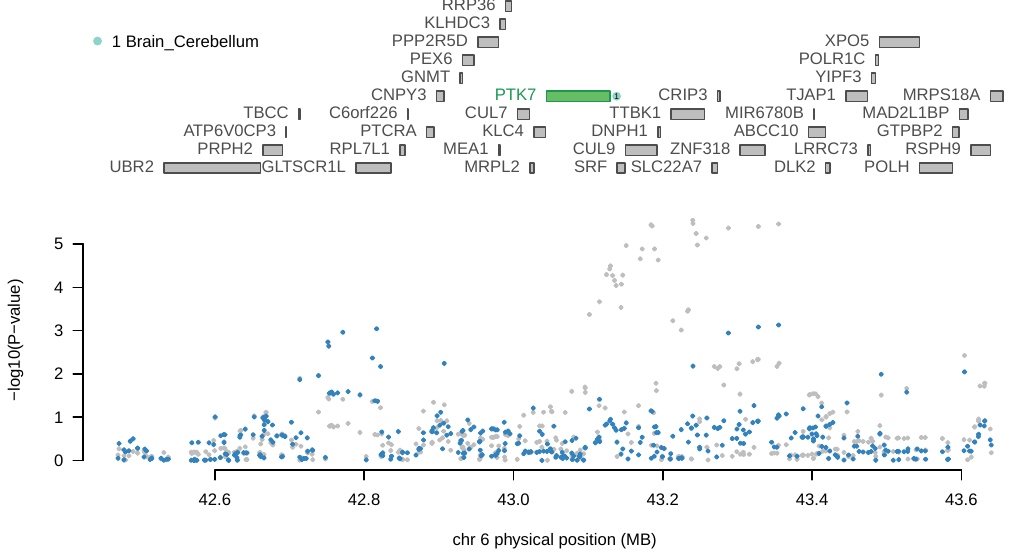

Figure S6g. Regional Association Plot for Locus 8 from g-factor Conditional Analyses. The top half of the panel displays all of the genes located in the locus window. Genes that were marginally significant are highlighted in blue while those that were jointly significant are highlighted in green. The color and numbering of the point next to jointly significant green genes indicates the specific tissue type (legend in upper-left) for which that gene was jointly significant. The bottom half depicts the Manhattan plot of the GWAS SNP effects within the locus prior to (grey) and after (blue) conditioning on the jointly significant genes in green. There were no marginally significant genome-wide effects within this locus after conditioning on the predicted expression of PTK7 gene in the cerebellum tissue type.

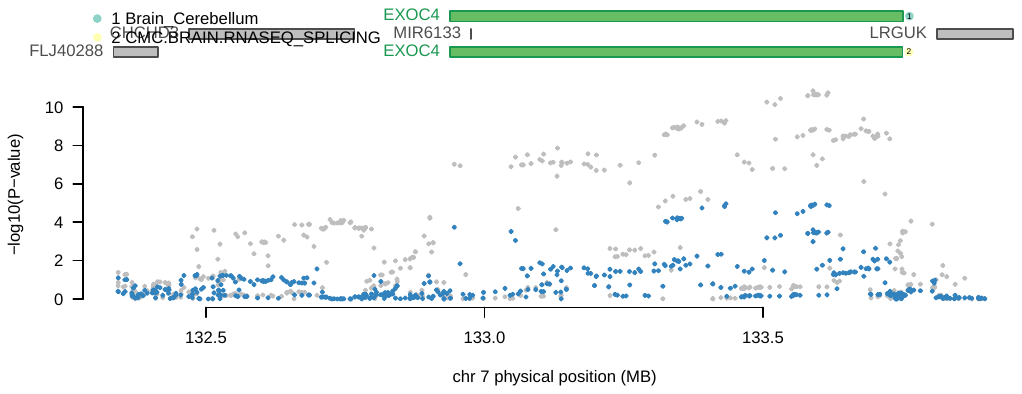

Figure S6h. Regional Association Plot for Locus 9 from g-factor Conditional Analyses. The top half of the panel displays all of the genes located in the locus window. Genes that were marginally significant are highlighted in blue while those that were jointly significant are highlighted in green. The color and numbering of the point next to jointly significant green genes indicates the specific tissue type (legend in upper-left) for which that gene was jointly significant. The bottom half depicts the Manhattan plot of the GWAS SNP effects within the locus prior to (grey) and after (blue) conditioning on the jointly significant genes in green. There were no marginally significant genome-wide effects within this locus after conditioning on the predicted expression of EXOC4 gene in both the cerebellum and Common Mind Consortium RNA-seq dlPFC tissue types.

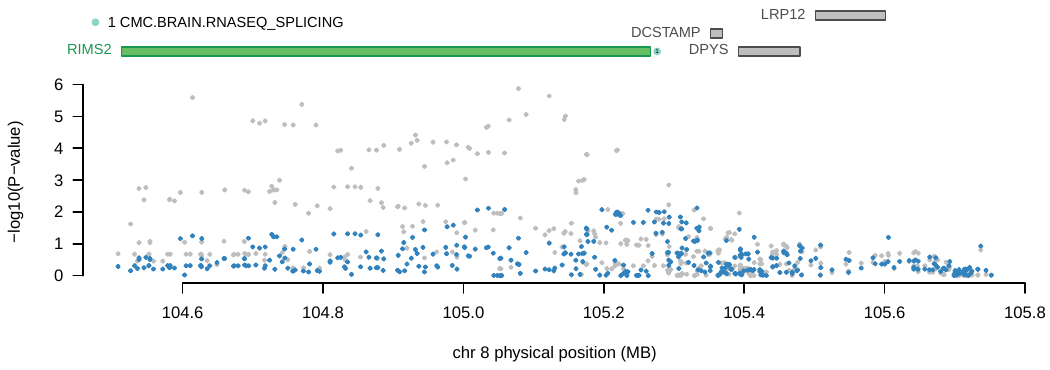

Figure S6i. Regional Association Plot for Locus 10 from g-factor Conditional Analyses. The top half of the panel displays all of the genes located in the locus window. Genes that were marginally significant are highlighted in blue while those that were jointly significant are highlighted in green. The color and numbering of the point next to jointly significant green genes indicates the specific tissue type (legend in upper-left) for which that gene was jointly significant. The bottom half depicts the Manhattan plot of the GWAS SNP effects within the locus prior to (grey) and after (blue) conditioning on the jointly significant genes in green. There were no marginally significant genome-wide effects within this locus after conditioning on the predicted expression of RIMS2 gene in the Common Mind Consortium RNA-seq dlPFC tissue type.

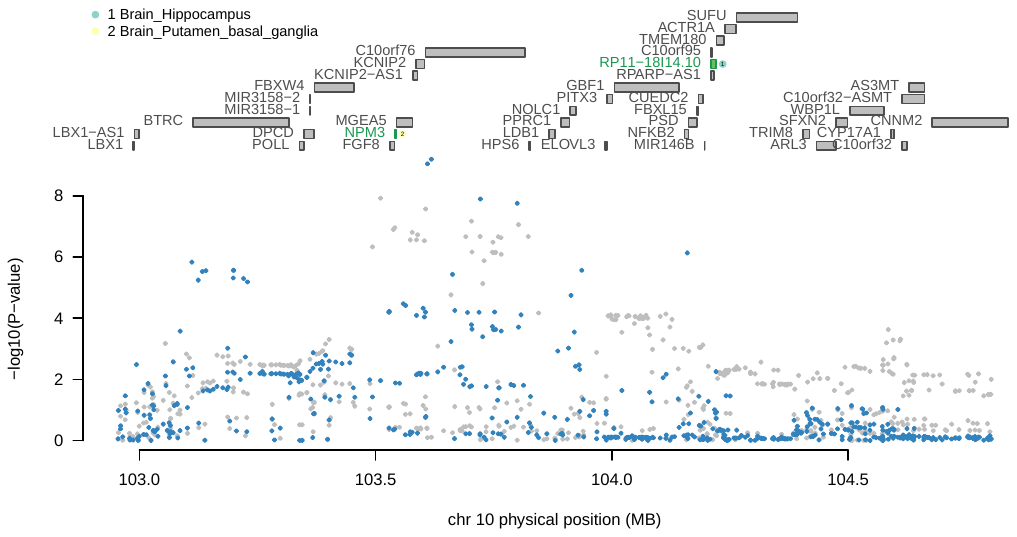
Figure S6j. Regional Association Plot for Locus 11 from g-factor Conditional Analyses. The top half of the panel displays all of the genes located in the locus window. Genes that were marginally significant are highlighted in blue while those that were jointly significant are highlighted in green. The color and numbering of the point next to jointly significant green genes indicates the specific tissue type (legend in upper-left) for which that gene was jointly significant. The bottom half depicts the Manhattan plot of the GWAS SNP effects within the locus prior to (grey) and after (blue) conditioning on the jointly significant genes in green. This particular locus reflects a unique case where some conditioned GWAS effects are more significant than unconditioned effects. This can occur when there is mismatch within a region between the LD reference and GWAS data.

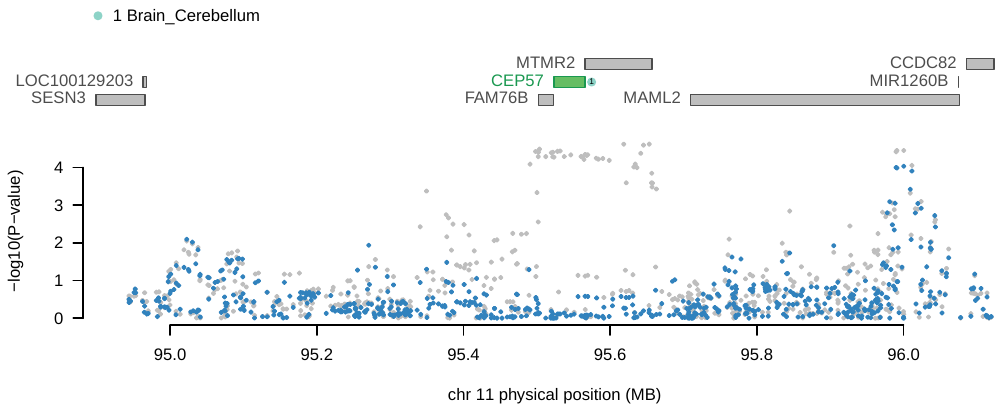

Figure S6k. Regional Association Plot for Locus 12 from g-factor Conditional Analyses. The top half of the panel displays all of the genes located in the locus window. Genes that were marginally significant are highlighted in blue while those that were jointly significant are highlighted in green. The color and numbering of the point next to jointly significant green genes indicates the specific tissue type (legend in upper-left) for which that gene was jointly significant. The bottom half depicts the Manhattan plot of the GWAS SNP effects within the locus prior to (grey) and after (blue) conditioning on the jointly significant genes in green. There were no marginally significant genome-wide effects within this locus after conditioning on the predicted expression of CEP57 gene in the cerebellum.

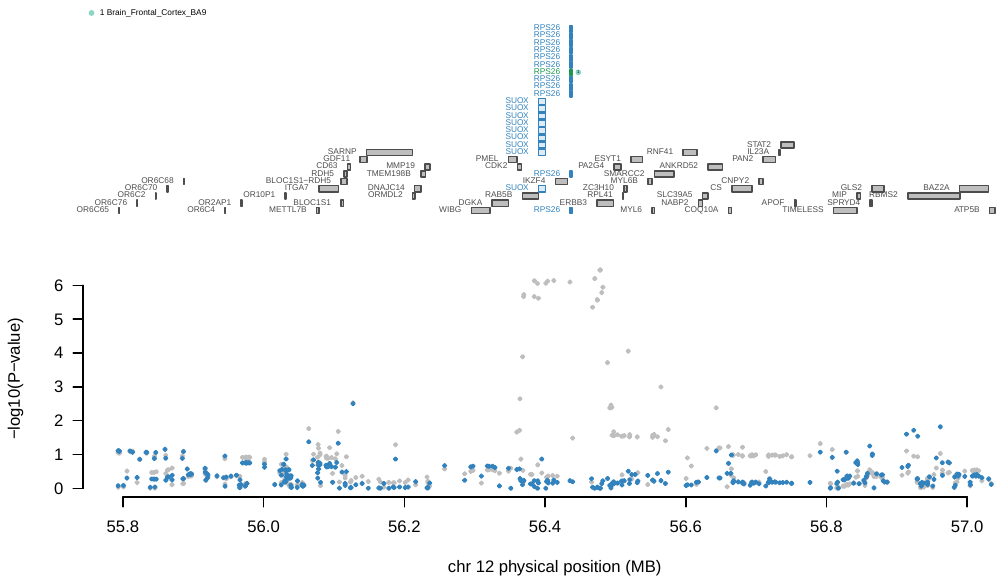

Figure S6l. Regional Association Plot for Locus 13 from g-factor Conditional Analyses. The top half of the panel displays all of the genes located in the locus window. Genes that were marginally significant are highlighted in blue while those that were jointly significant are highlighted in green. The color and numbering of the point next to jointly significant green genes indicates the specific tissue type (legend in upper-left) for which that gene was jointly significant. The bottom half depicts the Manhattan plot of the GWAS SNP effects within the locus prior to (grey) and after (blue) conditioning on the jointly significant genes in green. There were no marginally significant genome-wide effects within this locus after conditioning on the predicted expression of the RPS26 gene in the frontal cortex.

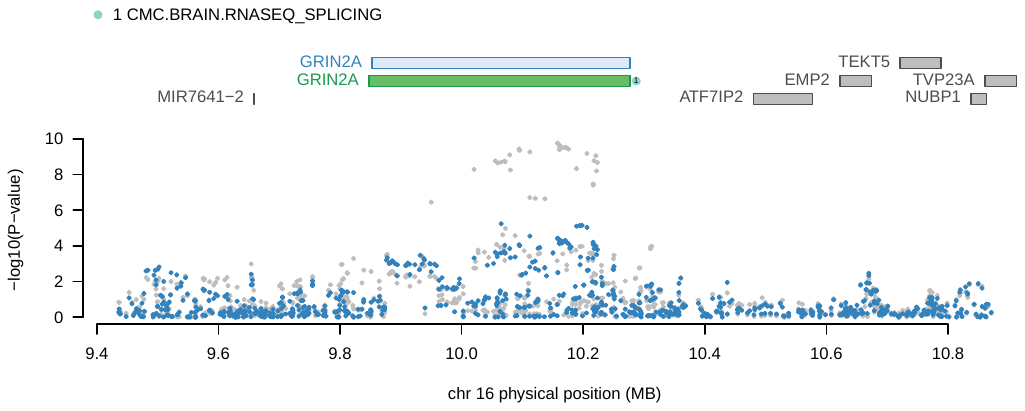

Figure S6m. Regional Association Plot for Locus 14 from g-factor Conditional Analyses. The top half of the panel displays all of the genes located in the locus window. Genes that were marginally significant are highlighted in blue while those that were jointly significant are highlighted in green. The color and numbering of the point next to jointly significant green genes indicates the specific tissue type (legend in upper-left) for which that gene was jointly significant. The bottom half depicts the Manhattan plot of the GWAS SNP effects within the locus prior to (grey) and after (blue) conditioning on the jointly significant genes in green. There were no significant genome-wide effects within this locus after conditioning on the predicted expression of the GRIN2A gene in the Common Mind Consortium RNA-seq dlPFC tissue.

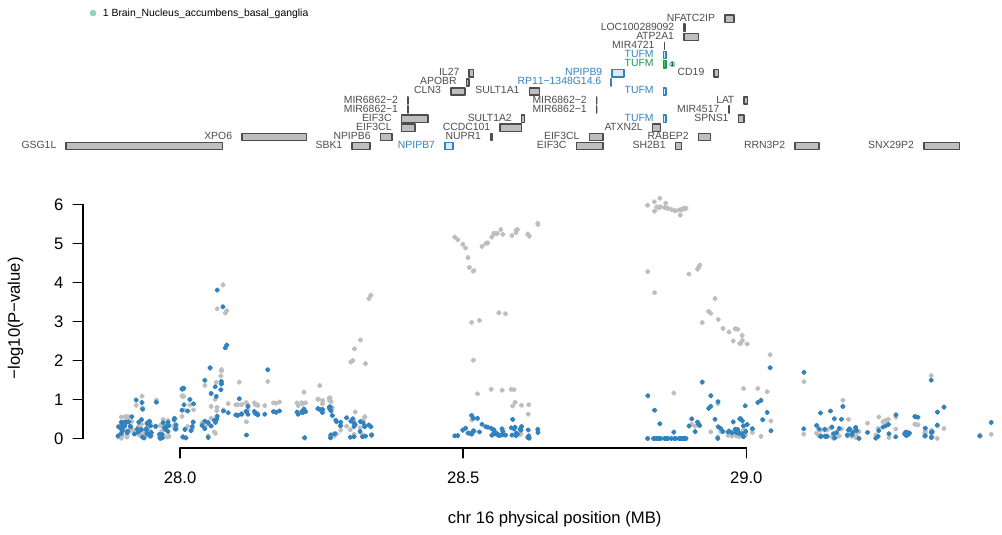

Figure S6n. Regional Association Plot for Locus 15 from g-factor Conditional Analyses. The top half of the panel displays all of the genes located in the locus window. Genes that were marginally significant are highlighted in blue while those that were jointly significant are highlighted in green. The color and numbering of the point next to jointly significant green genes indicates the specific tissue type (legend in upper-left) for which that gene was jointly significant. The bottom half depicts the Manhattan plot of the GWAS SNP effects within the locus prior to (grey) and after (blue) conditioning on the jointly significant genes in green. There were no marginally significant genome-wide effects within this locus after conditioning on the predicted expression of the TUFM gene in the nucleus accumbens.

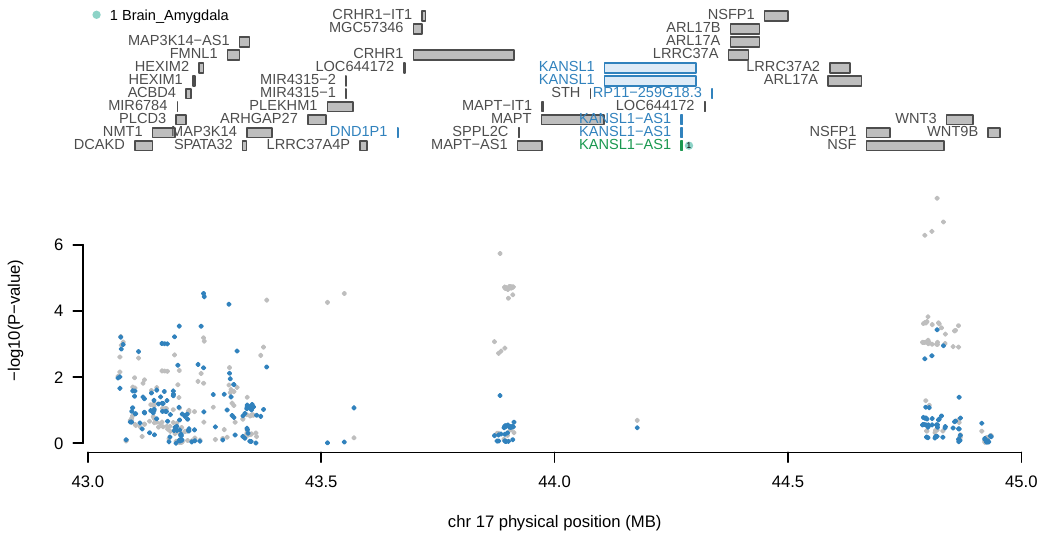

Figure S6o. Regional Association Plot for Locus 16 from g-factor Conditional Analyses. The top half of the panel displays all of the genes located in the locus window. Genes that were marginally significant are highlighted in blue while those that were jointly significant are highlighted in green. The color and numbering of the point next to jointly significant green genes indicates the specific tissue type (legend in upper-left) for which that gene was jointly significant. The bottom half depicts the Manhattan plot of the GWAS SNP effects within the locus prior to (grey) and after (blue) conditioning on the jointly significant genes in green. There were no marginally significant genome-wide effects within this locus after conditioning on the predicted expression of the KANSL1-AS1 gene in the amygdala.

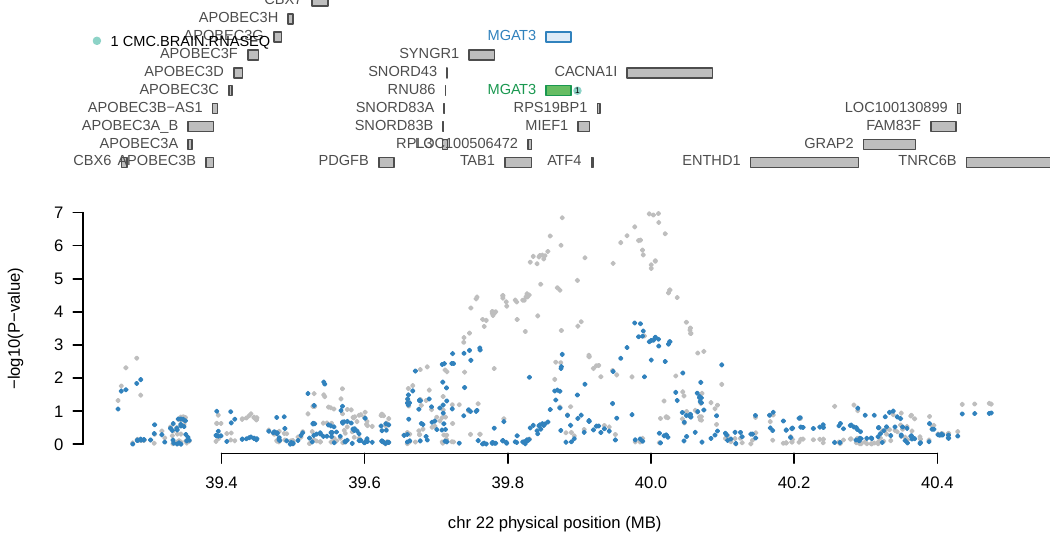

Figure S6p. Regional Association Plot for Locus 17 from g-factor Conditional Analyses. The top half of the panel displays all of the genes located in the locus window. Genes that were marginally significant are highlighted in blue while those that were jointly significant are highlighted in green. The color and numbering of the point next to jointly significant green genes indicates the specific tissue type (legend in upper-left) for which that gene was jointly significant. The bottom half depicts the Manhattan plot of the GWAS SNP effects within the locus prior to (grey) and after (blue) conditioning on the jointly significant genes in green. There were no marginally significant genome-wide effects within this locus after conditioning on the predicted expression of the MGAT3 gene in the Common Mind Consortium RNA-seq dlPFC tissue.

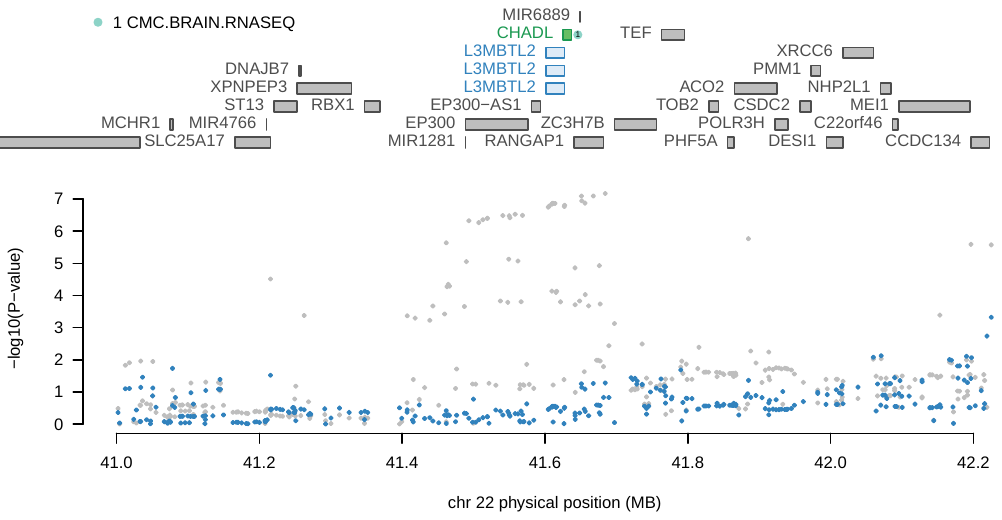

Figure S6q. Regional Association Plot for Locus 18 from g-factor Conditional Analyses. The top half of the panel displays all of the genes located in the locus window. Genes that were marginally significant are highlighted in blue while those that were jointly significant are highlighted in green. The color and numbering of the point next to jointly significant green genes indicates the specific tissue type (legend in upper-left) for which that gene was jointly significant. The bottom half depicts the Manhattan plot of the GWAS SNP effects within the locus prior to (grey) and after (blue) conditioning on the jointly significant genes in green. There were no marginally significant genome-wide effects within this locus after conditioning on the predicted expression of the CHADL gene in the Common Mind Consortium RNA-seq dlPFC tissue.

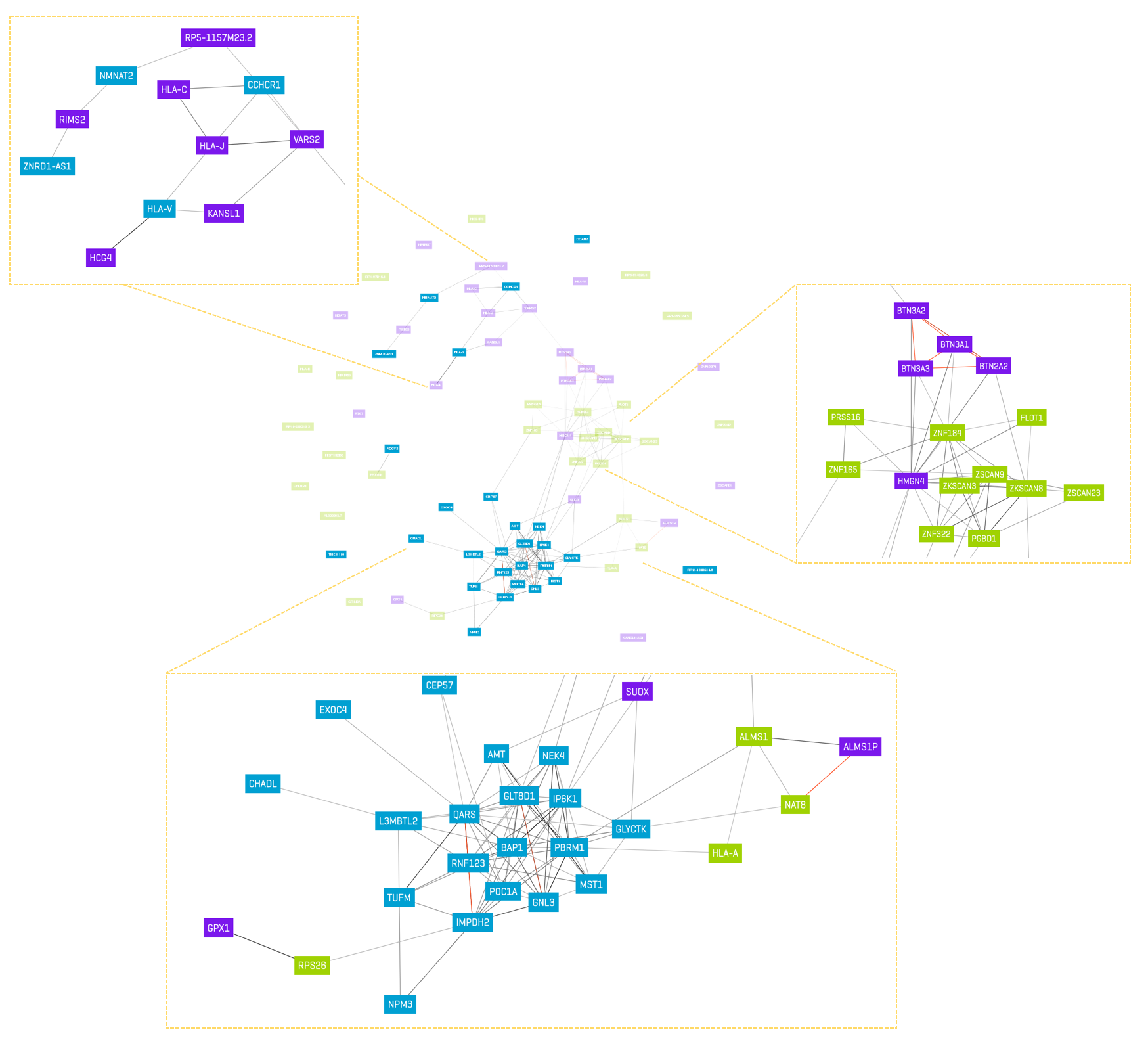

Figure S7. g-factor Gene Co-expression Network. Figure depicts gene co-expression network for g-factor created using *Gene Network v2.0* for N=31,499 public RNA sequencing samples. Genes used as input to create the co-expression network included those genes that were significant at a Bonferroni corrected threshold for 52,849 tests and did not overlap with significant Q_Gene_ hits for the same gene and tissue type. These genes were restricted still further to those unique gene IDs across tissue types for a total of 79 genes used as input. As 3 of these genes were not in the gene network database, the plot above was constructed using a total of 76 gene IDs as input. Gene cluster 1 is depicted in blue (27 total genes). Gene cluster 2 is depicted in green (27 total genes). Gene cluster 3 is depicted in purple (22 total genes). Darker lines indicate stronger co-expression, with red lines depicted negative patterns of co-expression. Due to the size of the network, particular clusters of genes are depicted in greater detail in the yellow dashed boxes.

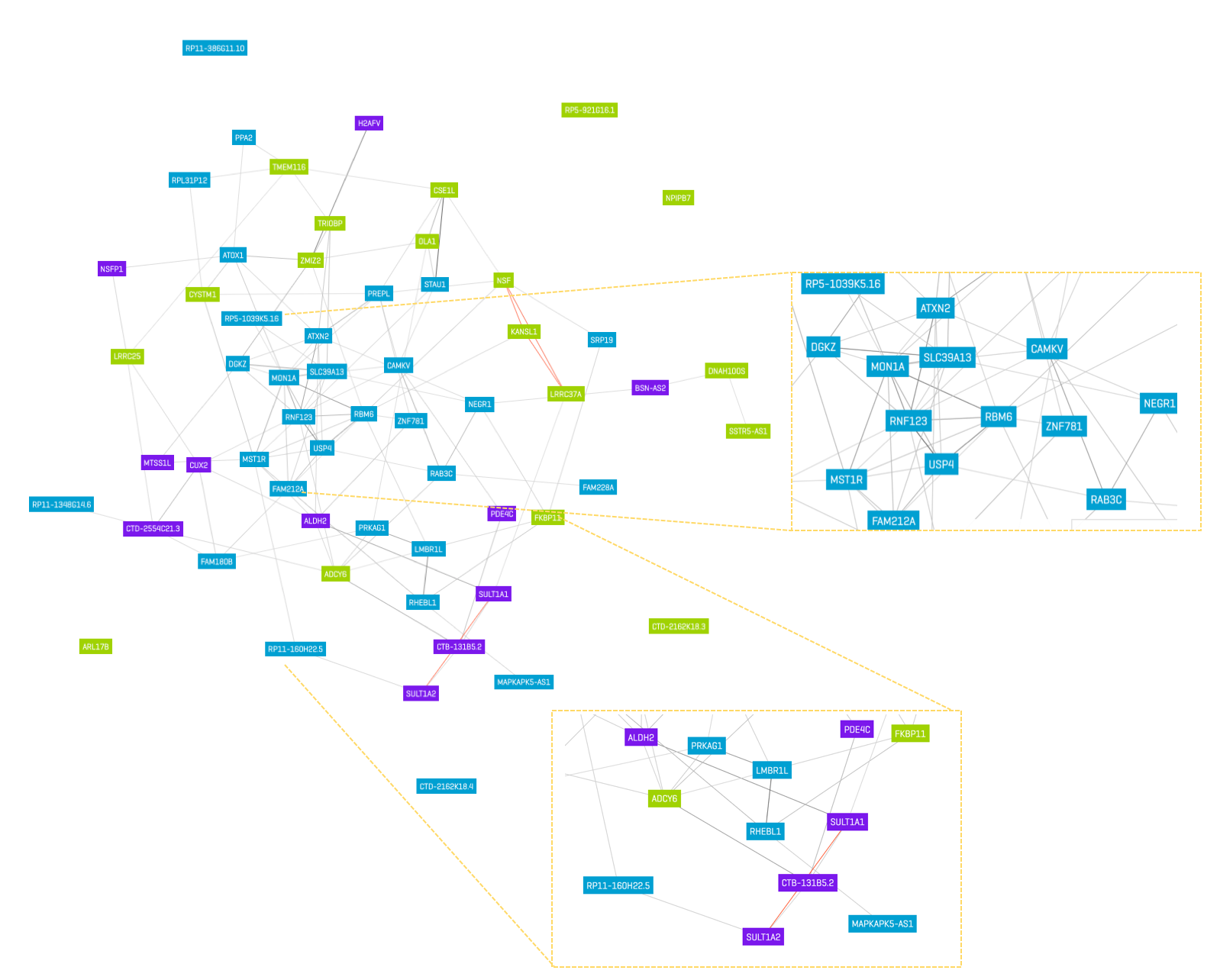

Figure S8. Q_Gene_ Gene Co-expression Network. Figure depicts gene co-expression network for Q_Gene_ created using *Gene Network v2.0* for N=31,499 public RNA sequencing samples. Genes used as input to create the co-expression network included the 62 unique gene IDs that were significant at a Bonferroni corrected threshold for 52,849 tests. As 3 of these genes were not in the gene network database, the plot above was constructed using a total of 59 gene IDs as input. Gene cluster 1 is depicted in blue (30 total genes). Gene cluster 2 is depicted in green (18 total genes). Gene cluster 3 is depicted in purple (11 total genes). Darker lines indicate stronger co-expression, with red lines depicted negative patterns of co-expression. Due to the size of the network, particular clusters of genes are depicted in greater detail in the yellow dashed boxes.

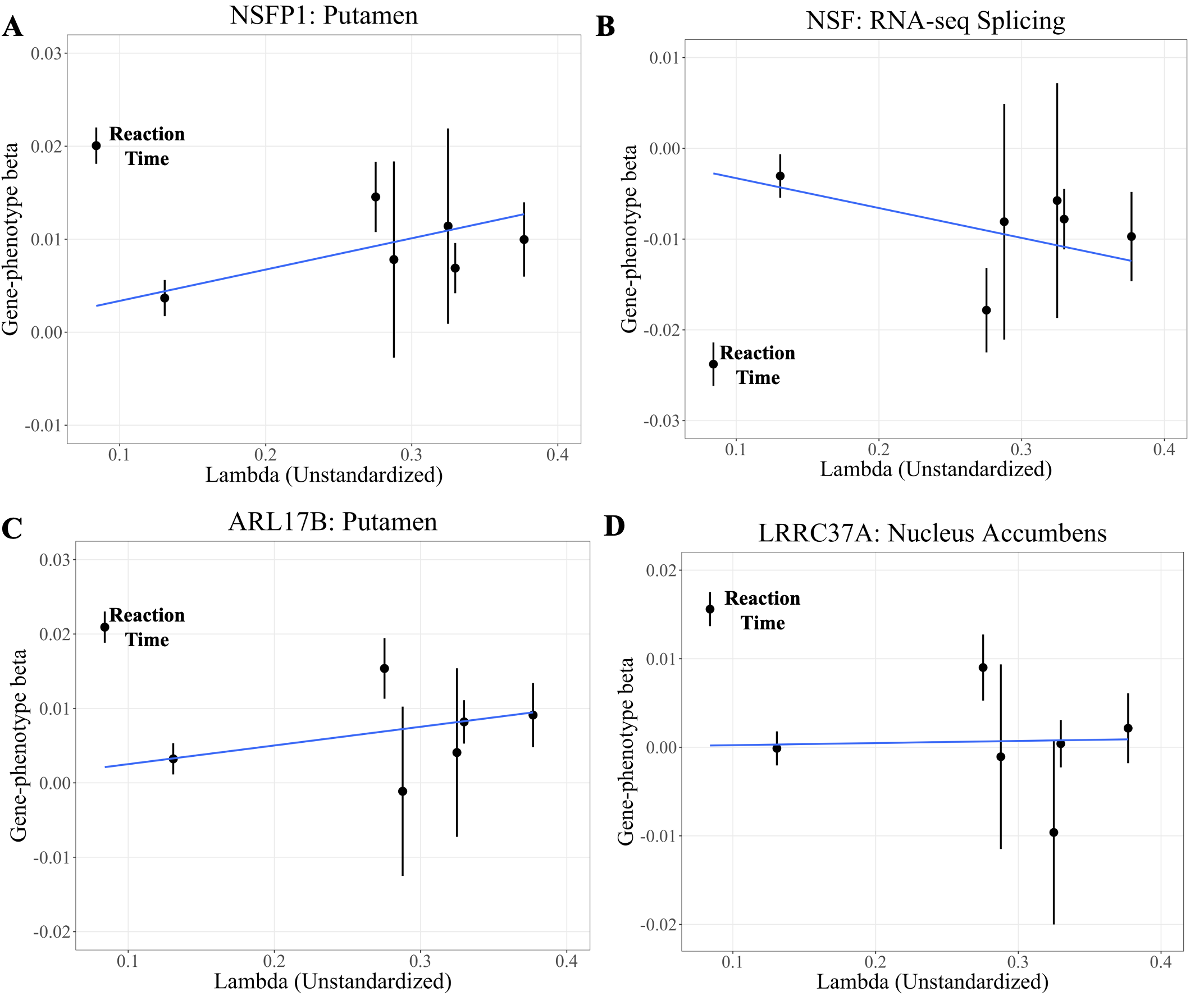

Figure S9. Scatterplot of 17q21.31 Q_Gene_ hits. Scatter plot of Gene-phenotype regression coefficients (betas) estimated from FUSION against unstandardized genetic factor loadings form common factor model for genomic *g* estimated using LDSC genetic covariance matrix as input. Scatter plots are depicted for the 4 unique Q_Gene_ hits for the most significant tissue in the 17q21.31 region. Panel A depicts the NSFP1 gene in the putamen panel. Panel B depicts the NSF gene in the RNA-seq splicing panel. Panel C depicts the ARL17B gene in the putamen panel. Panel D depicts the LRRC37A gene in the nucleus accumbens panel. Error bars reflect +/- 1 standard error of the betas. The solid blue line reflects the linear regression line based on all seven data points with the intercepts fixed to 0 to reflect the expectation from a common pathways model that the gene-phenotype regression relationship is 0 for an indicator that loads on the factor at 0. The data point for reaction time is highlighted across panels as it deviated the most strongly from this regression line.

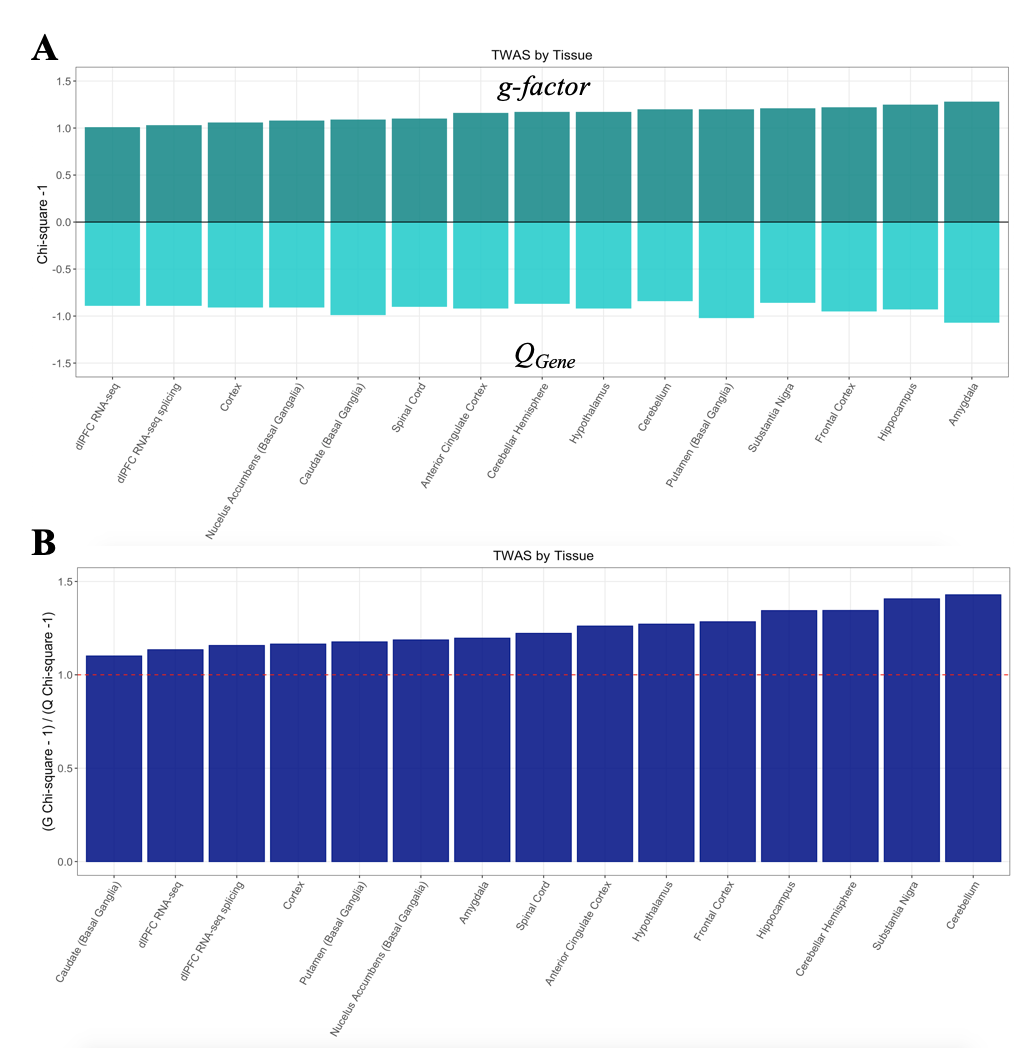

Figure S10. Mean **χ^2^** across Tissues. Panel A depicts the for the g-factor on the top half and Q_Gene_*-1 on the bottom for the mean χ^2^ – 1. Bars are depicted in ascending order of the average values for the g-factor. Panel B depicts the ratio of mean χ^2^ – 1 for the g-factor over Q_Gene_ and are again depicted in ascending order for this particular ratio. A red dashed line is depicted at 1, as this would indicate equal signal for Q_Gene_ and the *g*-factor. For both panels, Q_Gene_ is scaled to a 1 degree of freedom χ^2^ test statistic for comparative purposes, and 1 is subtracted from all χ^2^ averages given a χ^2^ null of 1.

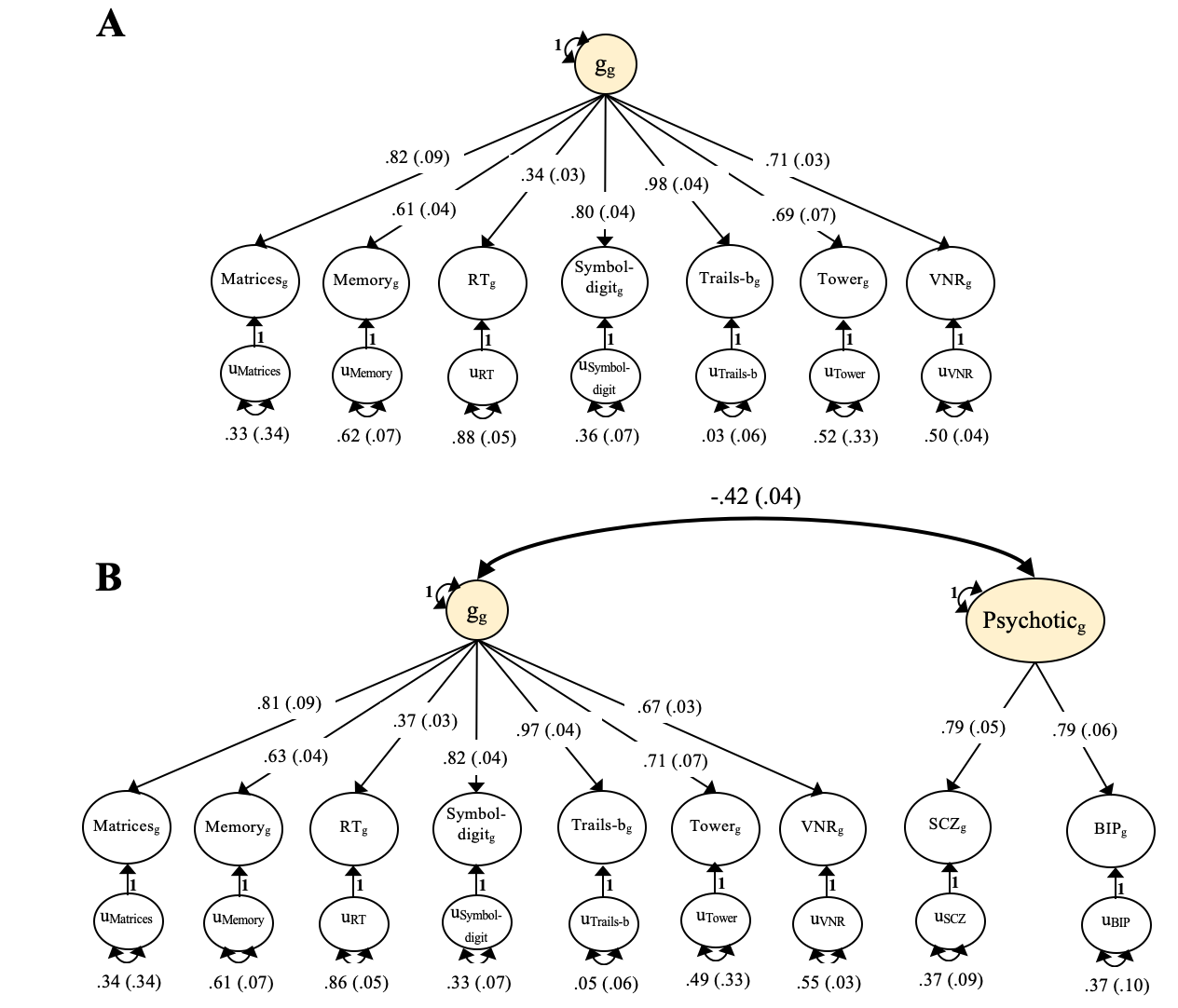

Figure S11. Factor Models estimated using Genome-wide, Zero-order Matrix. *Panel A:* Standardized results for g-factor model fit to the genome-wide S-LDSC matrix. This reflects the baseline model used to estimate enrichment of genetic variance of the g-factor, where all factor loadings are fixed from this model in Step 1 and the indicator uniquenesses (i.e., residual variances) and factor variance are re-estimated within each functional annotation. The factor loadings will differ very slightly from those presented in the original description of genomic g^1^, as those estimates were obtained from a common factor model fit to a matrix estimated using LDSC (as opposed to S-LDSC). *Panel B:* Standardized results for correlated factors model between a g-factor and psychotic disorders factor fit to S-LDSC matrix for the annotation including all SNPs. This model was used to examine enrichment of the genetic correlation between these two factors. For identification purposes, the factor loadings of the psychotic disorders are set to equality across schizophrenia and bipolar disorder. Indicators in both panels are presented as circles to reflect the fact that these are unobserved heritability estimates from S-LDSC.

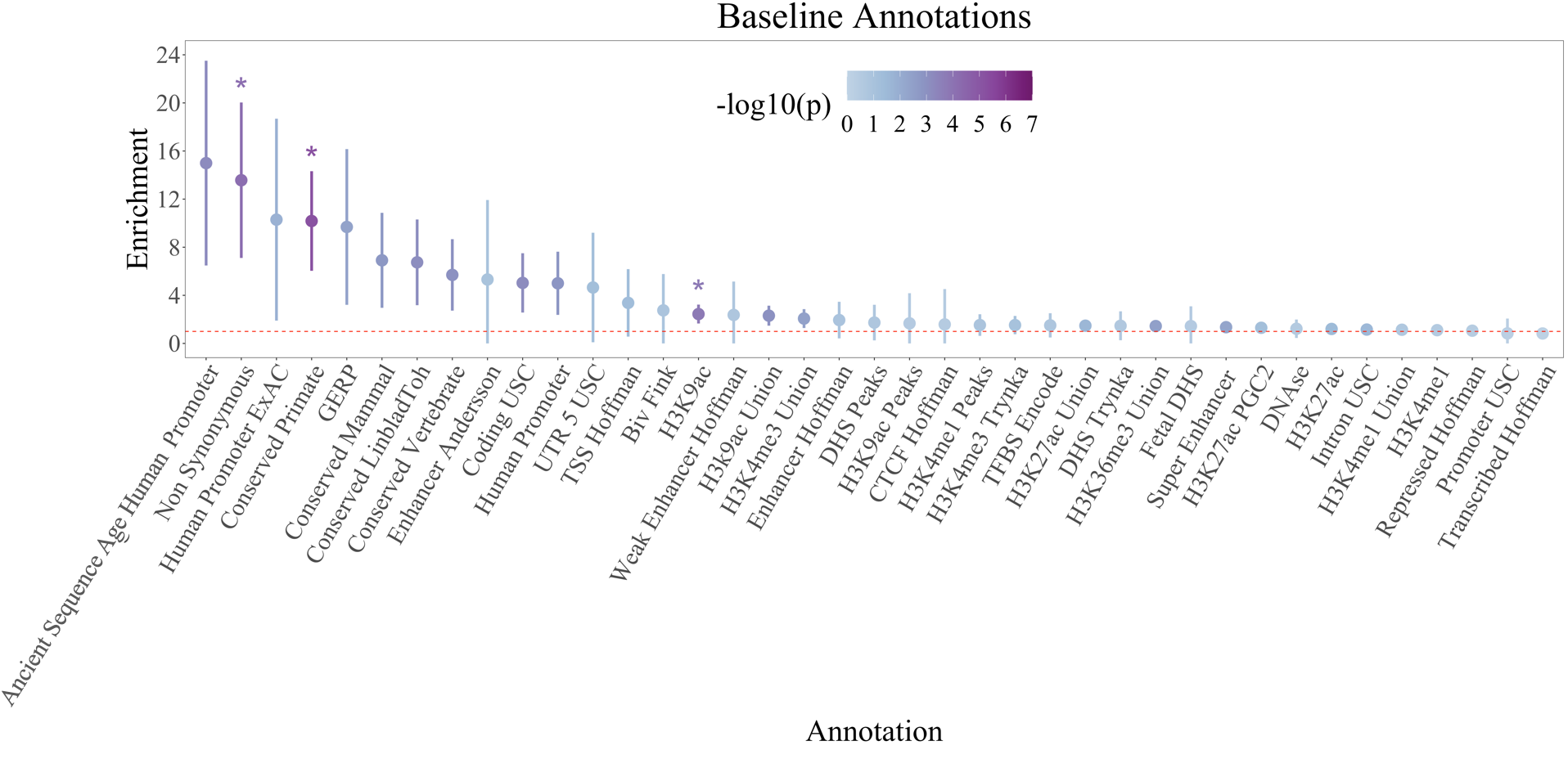

Figure S12a. Enrichment of Baseline Annotations for g-factor. Dots are depicted in descending order based on the point estimate for enrichment of the *g-*factor. Dots are shaded according to the significance of the enrichment estimate and dots that were significant at a Bonferroni corrected threshold for 155 tests are depicted with a *. The red dashed line reflects the null (enrichment = 1). Error bars depict 95% CIs. The scaling of the y-axis across enrichment graphs differs due to discrepant ranges in CIs across annotations.

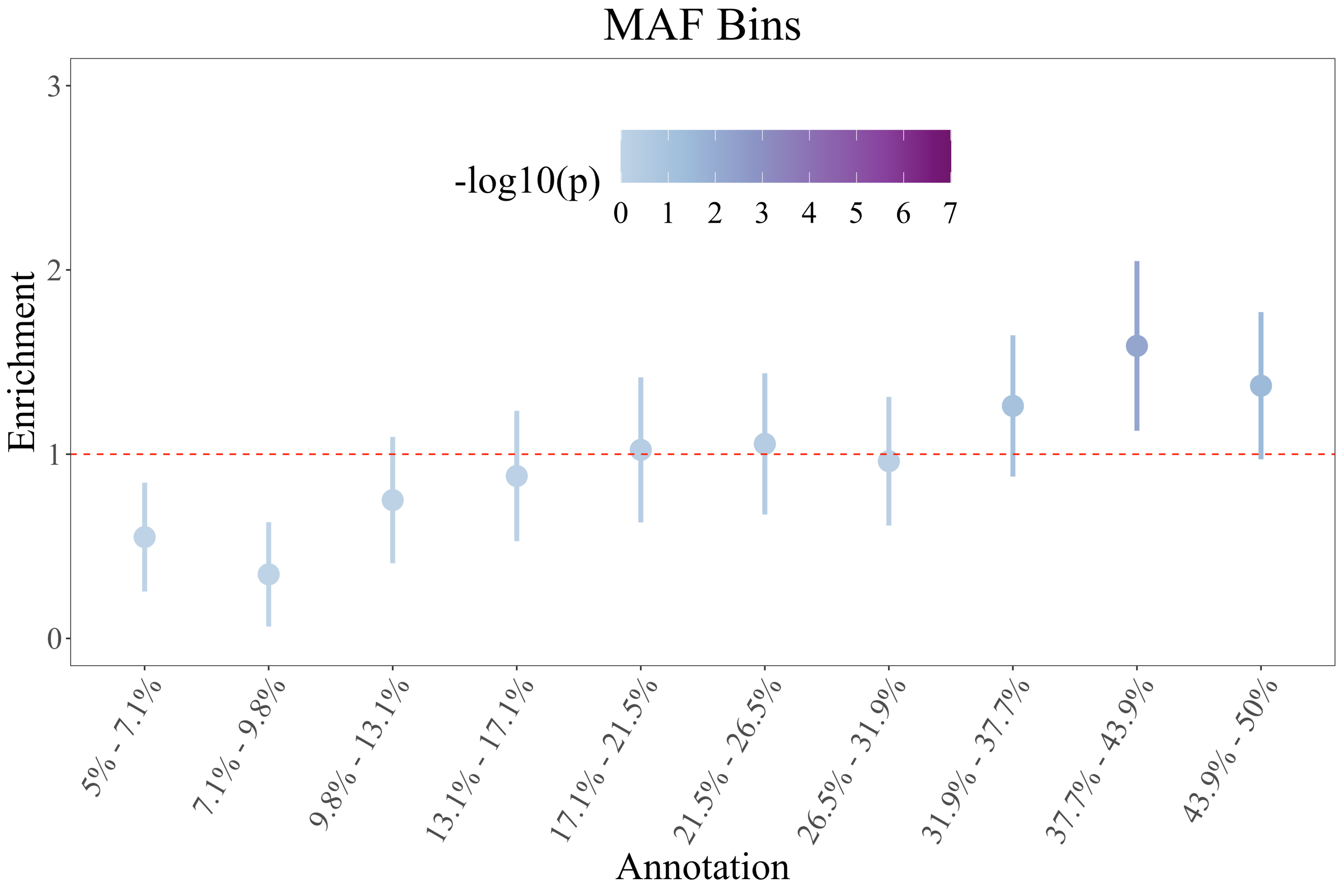

Figure S12b. Enrichment of MAF Annotations for g-factor. Dots are depicted in order of the minor allele frequency bins. Dots are shaded according to the significance of the enrichment estimate. No MAF bins were significant at a Bonferroni corrected threshold for 155 tests. The red dashed line reflects the null (enrichment = 1). Error bars depict 95% CIs. The scaling of the y-axis across enrichment graphs differs due to discrepant ranges in CIs across annotations.

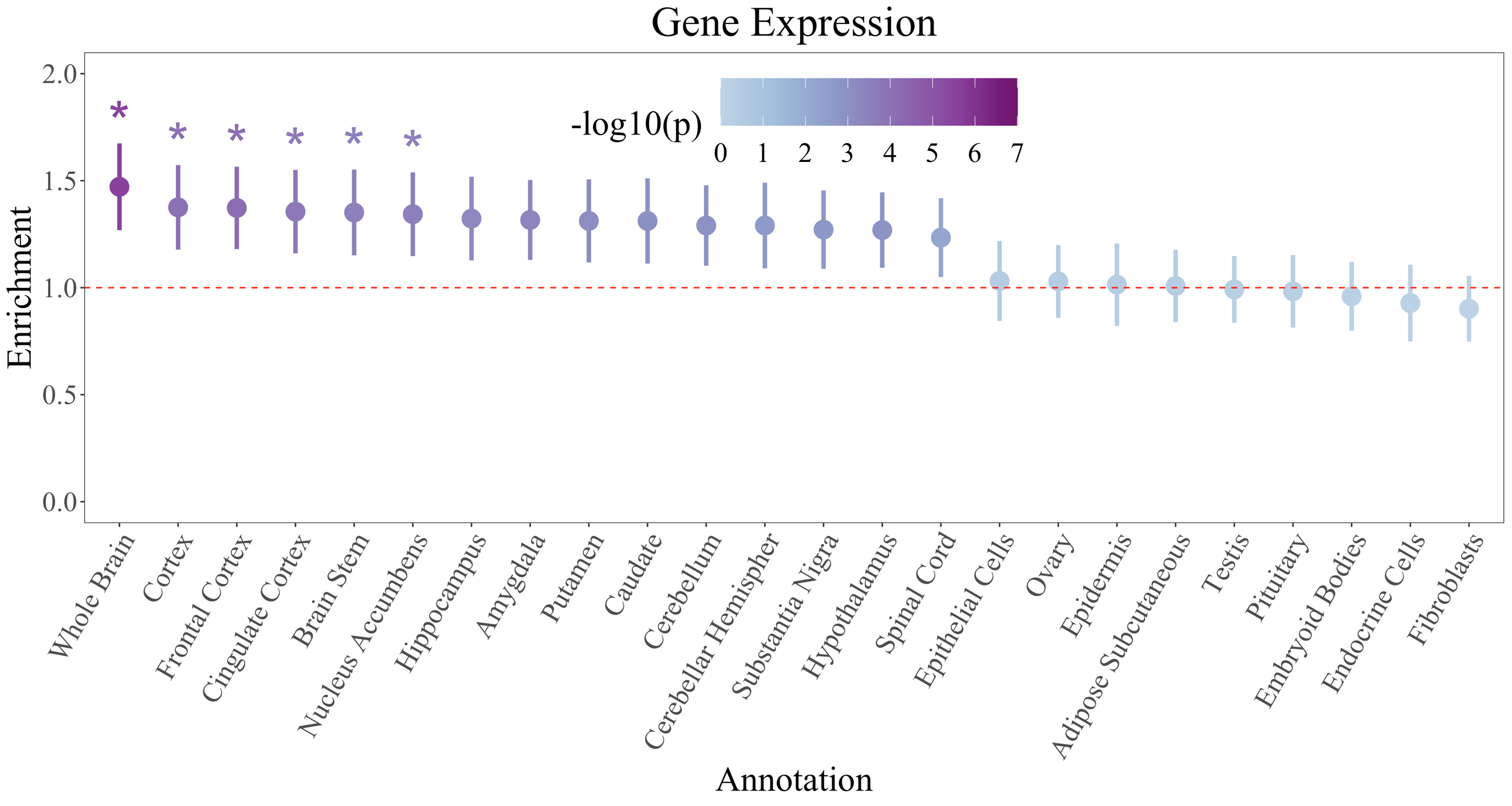

Figure S12c. Enrichment of Gene Expression Annotations for g-factor. Dots are depicted in descending order based on the point estimate for enrichment of the *g-*factor. Dots are shaded according to the significance of the enrichment estimate, with dots that were significant at a Bonferroni corrected threshold for 155 tests depicted with a *. The red dashed line reflects the null (enrichment = 1). Error bars depict 95% CIs. The scaling of the y-axis across enrichment graphs differs due to discrepant ranges in CIs across annotations.

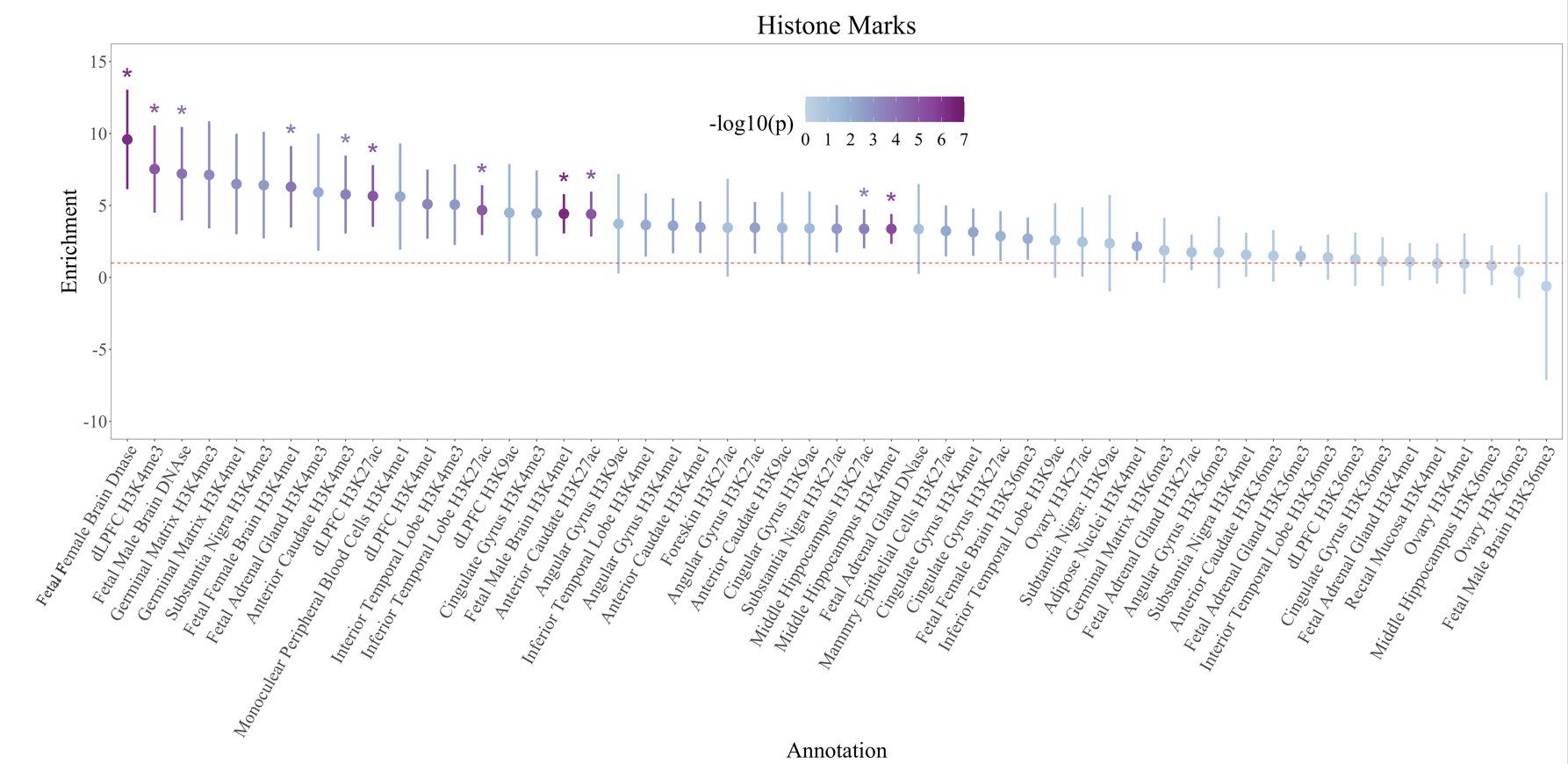

Figure S12d. Enrichment of Histone Mark Annotations for g-factor. Dots are depicted in descending order based on the point estimate for enrichment of the *g-*factor. Dots are shaded according to the significance of the enrichment estimate, with dots that were significant at a Bonferroni corrected threshold for 155 tests depicted with a *. The red dashed line reflects the null (enrichment = 1). Error bars depict 95% CIs. The scaling of the y-axis across enrichment graphs differs due to discrepant ranges in CIs across annotations.

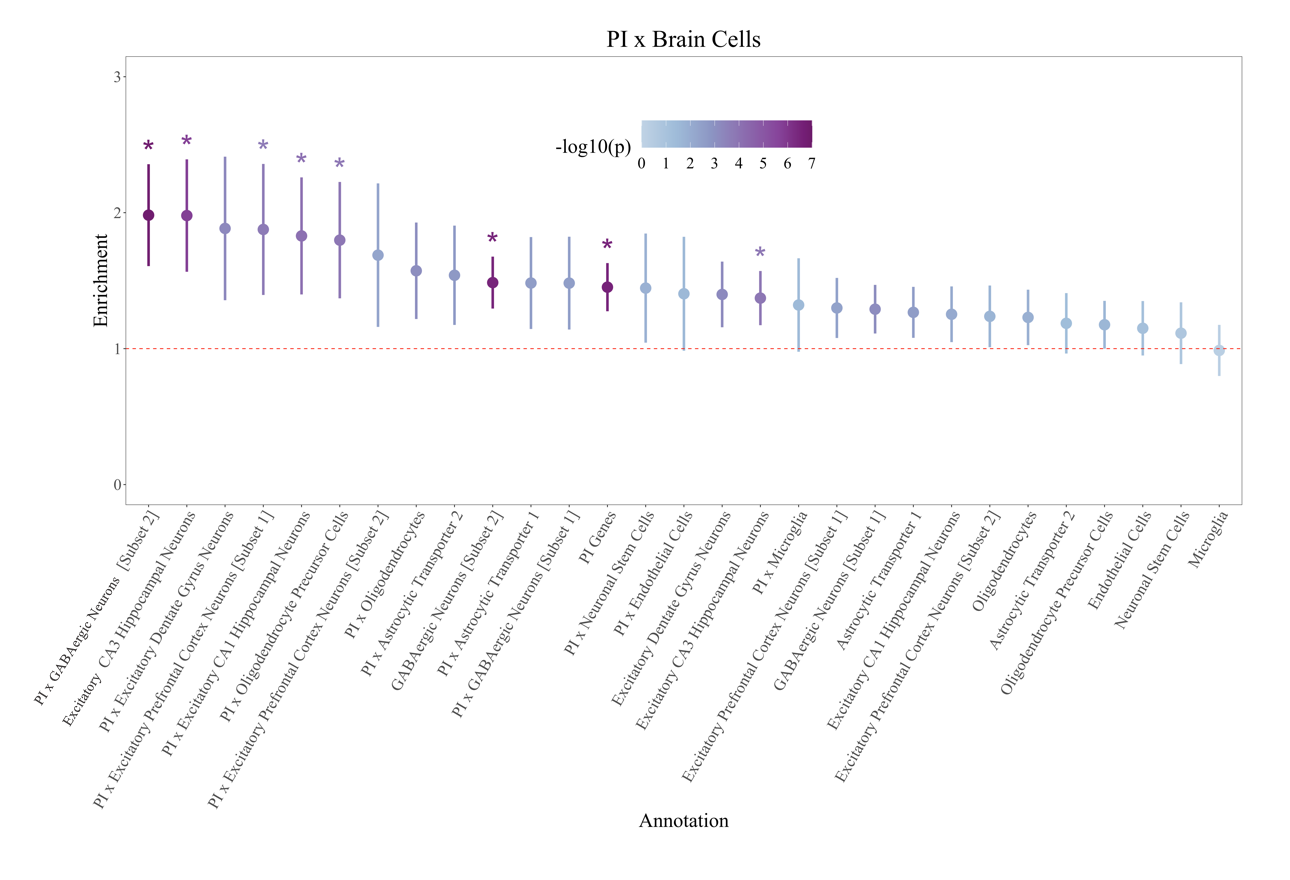

Figure S12e. Enrichment of PI x Brain Cell Annotations for g-factor. Dots are depicted in descending order based on the point estimate for enrichment of the *g-*factor. Dots are shaded according to the significance of the enrichment estimate. The red dashed line reflects the null (enrichment = 1), error bars depict 95% CIs, and dots that were significant at a Bonferroni corrected threshold for 155 tests are depicted with a *.

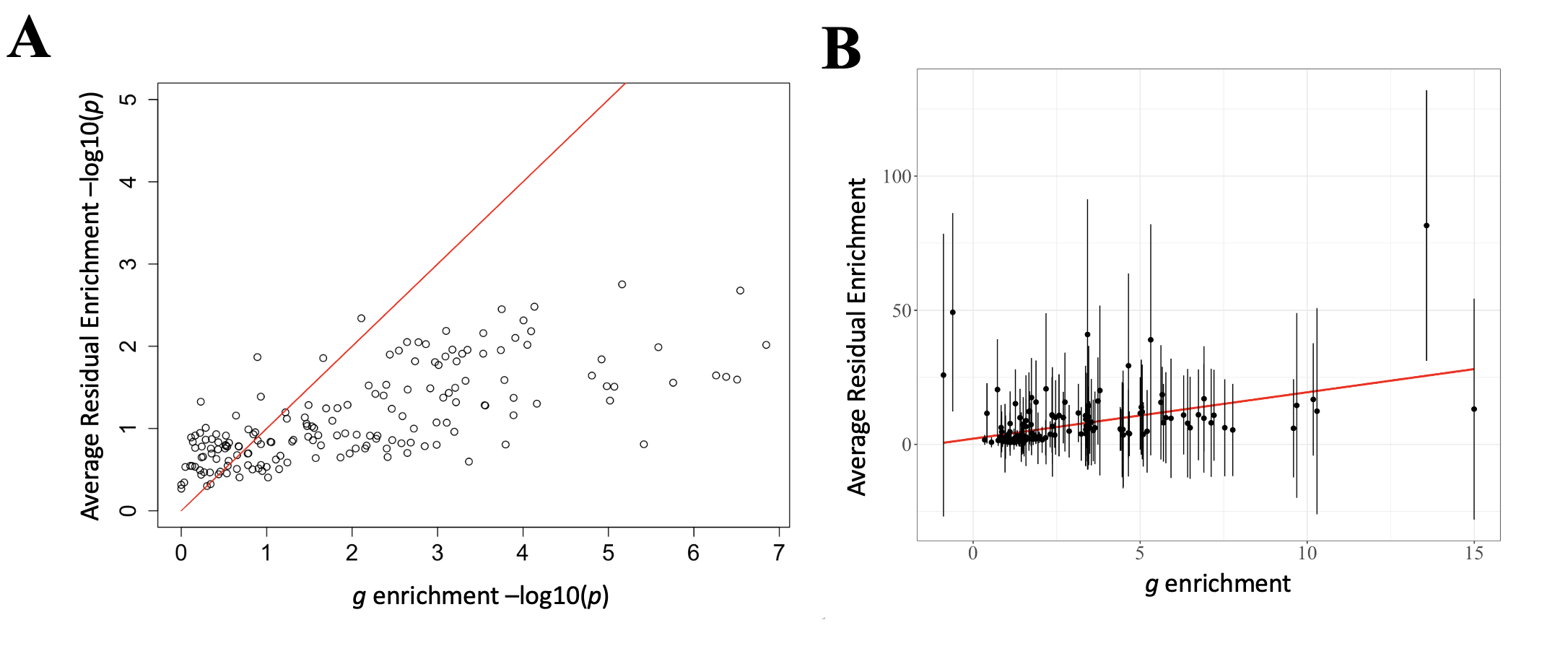

Figure S13. Scatterplots of residual and genetic *g* enrichment. *Panel A* depicts the relationship between the -log10 p-values for *g* on the x-axis and the average of the -log10 p-values across the residuals for the seven cognitive indicators on the y-axis. *Panel B* depicts the same scatter plot but with enrichment point estimates on both axes. Error bars reflect +/- 1 SE for the residual point estimates. Red lines reflect the g enrichment estimates predicting itself in both panels, with dots below the line reflecting more significant estimates for g relative to the residuals.

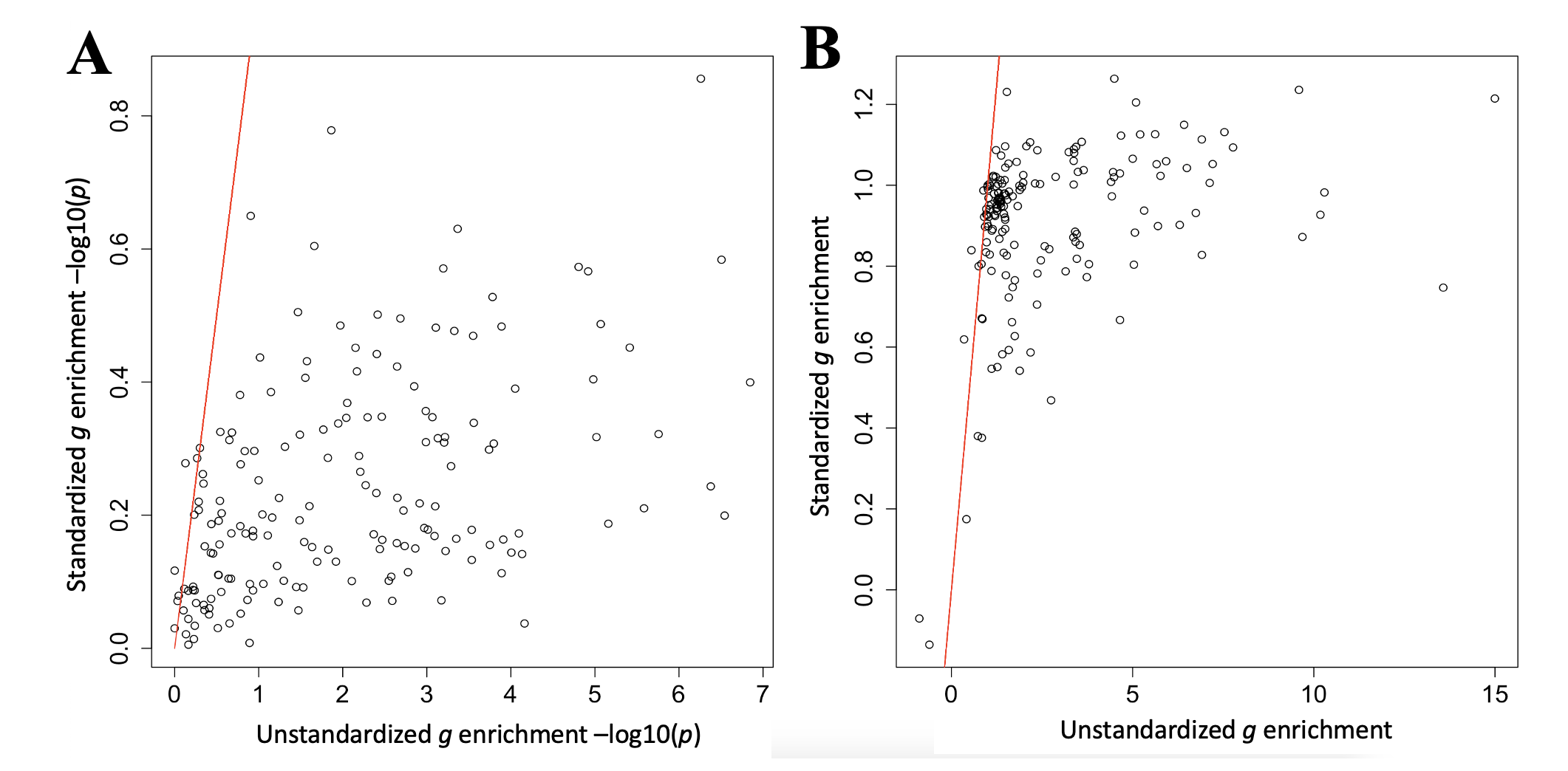

Figure S14. Scatter plots for *g* enrichment fit to stratified covariance and correlation matrices. *Panel A* depicts the relationship between the -log10 p-values for *g* fit to stratified covariance matrices (unstandardized) on the x-axis and to stratified correlation matrices (standardized) on the y-axis. *Panel B* depicts the same scatter plot but with enrichment point estimates on both axes. Red lines reflect the unstandardized g enrichment estimates predicting itself in both panels, with dots below the line reflecting more significant estimates for unstandardized *g* relative to standardized *g*.

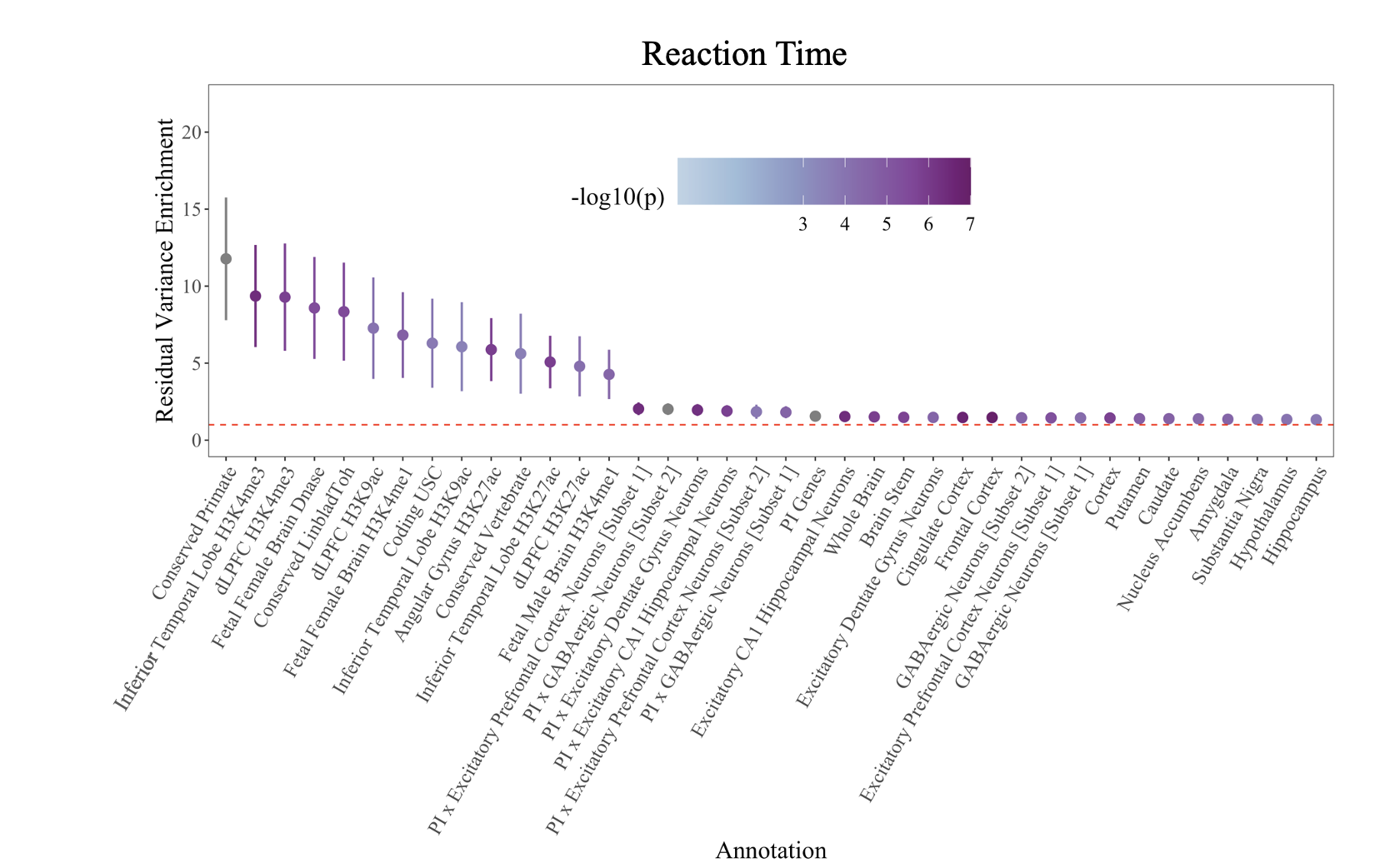

Figure S15a. Enrichment for Residual Variance in Reaction Time. Dots are depicted in descending order based on the point estimate for enrichment of the residual variance in reaction time. Figure depicts the 38 significant estimates for reaction time. Dots are shaded according to the significance of the enrichment estimate. The red dashed line reflects the null (enrichment = 1) and error bars depict 95% CIs.

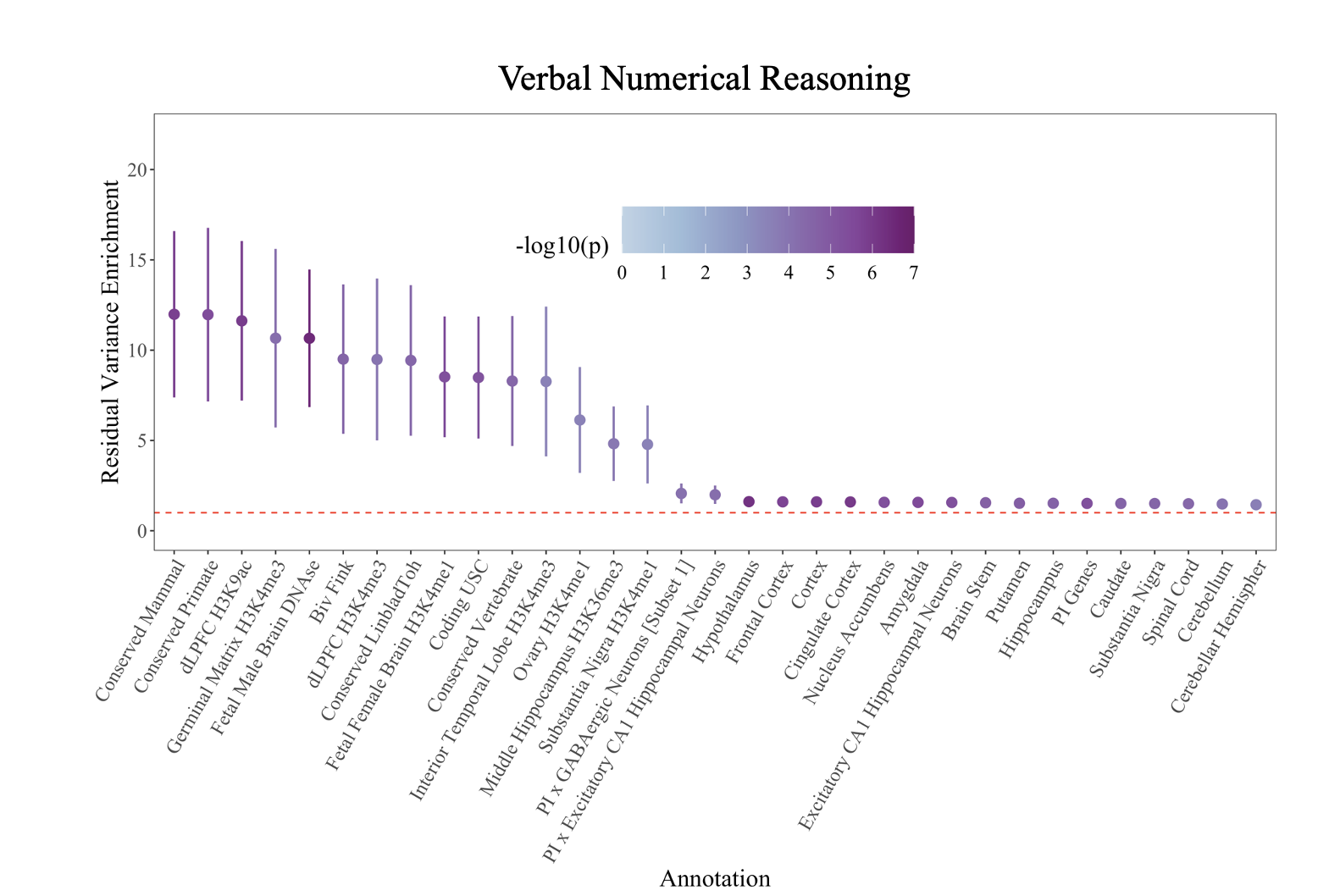

Figure S15b. Enrichment for Residual Variance in Verbal Numerical Reasoning. Dots are depicted in descending order based on the point estimate for enrichment of the residual variance in verbal numerical reasoning. Figure depicts the 33 significant estimates for verbal numerical reasoning. Dots are shaded according to the significance of the enrichment estimate. The red dashed line reflects the null (enrichment = 1) and error bars depict 95% CIs.

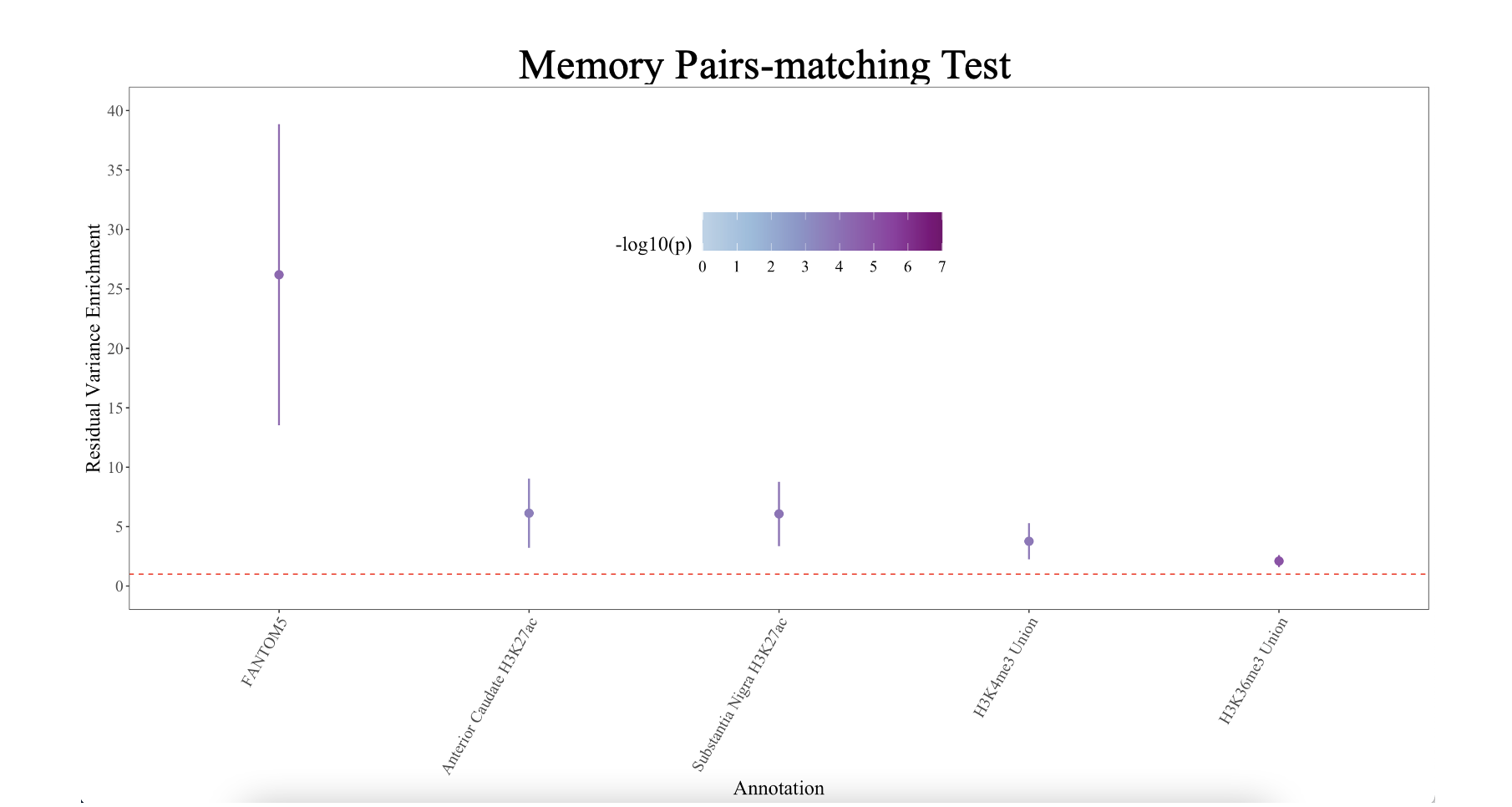

Figure S15c. Enrichment for Residual Variance in the Memory Pairs-matching Test. Dots are depicted in descending order based on the point estimate for enrichment of the residual variance in the memory pairs-matching test. Figure depicts the 5 significant estimates for memory pairs-matching test. Dots are shaded according to the significance of the enrichment estimate. The red dashed line reflects the null (enrichment = 1) and error bars depict 95% CIs.

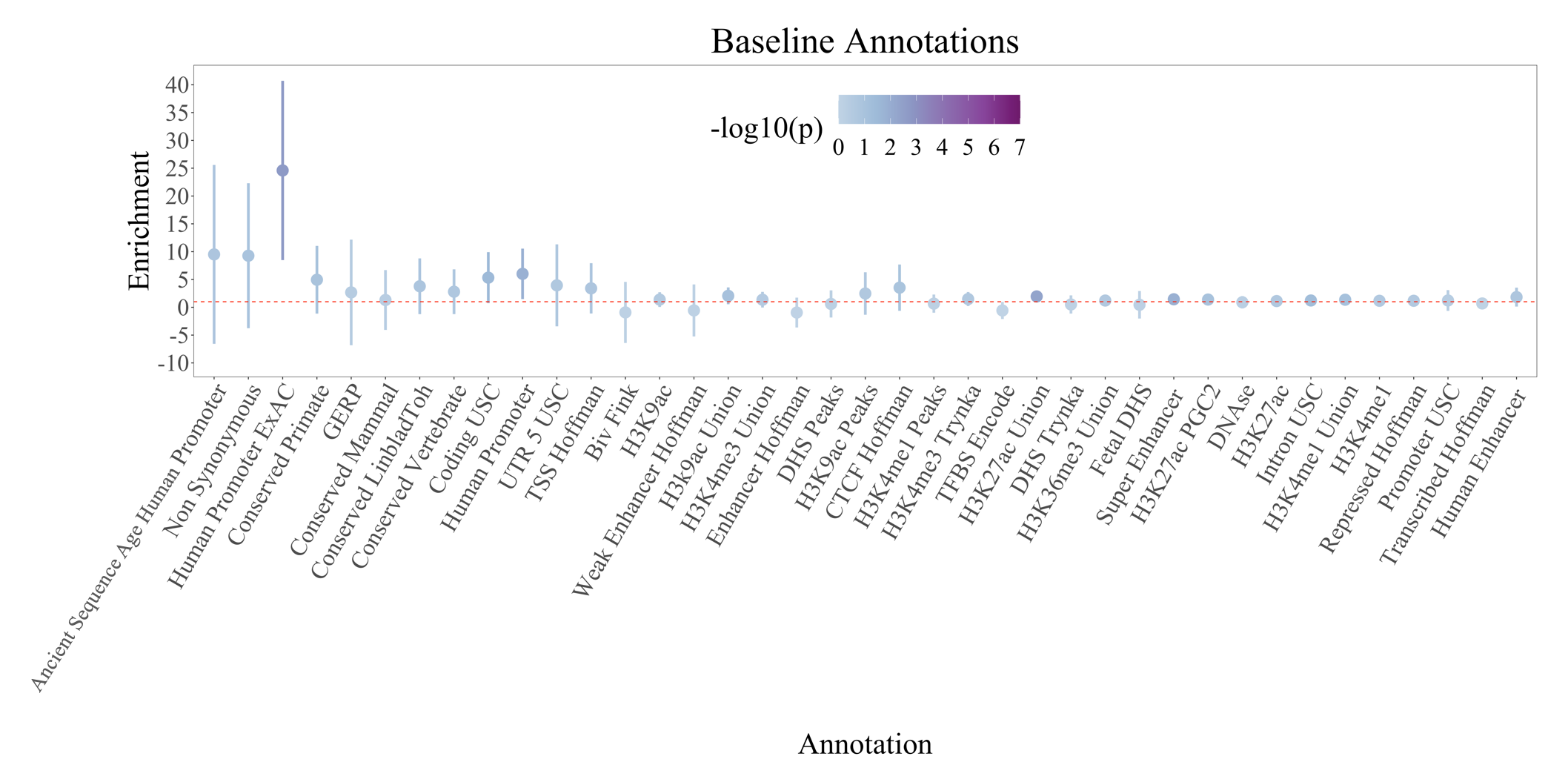

Figure S16a. Enrichment of Baseline Annotations for covariance between the *g*-factor and psychotic disorders factor. For comparative purposes, dots are depicted in descending order based on the point estimate for enrichment of the *g-*factor as per Figure S6a. Dots are shaded according to the significance of the enrichment estimate. No baseline annotations were at a Bonferroni corrected threshold for 155 tests. The red dashed line reflects the null (enrichment = 1). Error bars depict 95% CIs. The scaling of the y-axis differs across enrichment graphs due to widely discrepant ranges in point estimates and CIs across annotations.

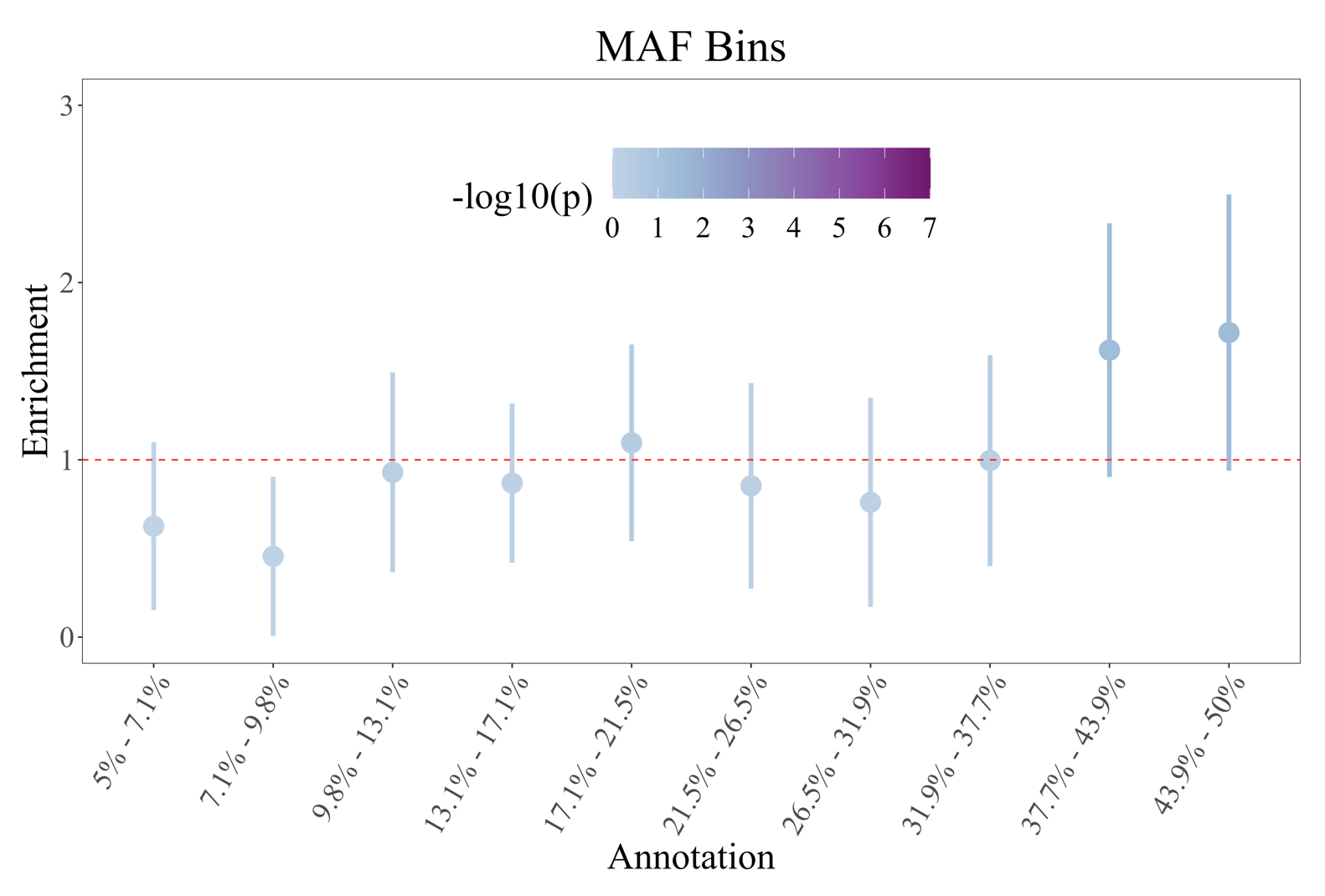

Figure S16b. Enrichment of MAF Annotations for covariance between the *g*-factor and psychotic disorders factor. Dots are depicted in order of the minor allele frequency bins. Dots are shaded according to the significance of the enrichment estimate. No MAF bins were significant at a Bonferroni corrected threshold for 155 tests. The red dashed line reflects the null (enrichment = 1). Error bars depict 95% CIs. The scaling of the y-axis across enrichment graphs differs due to discrepant ranges in CIs across annotations.

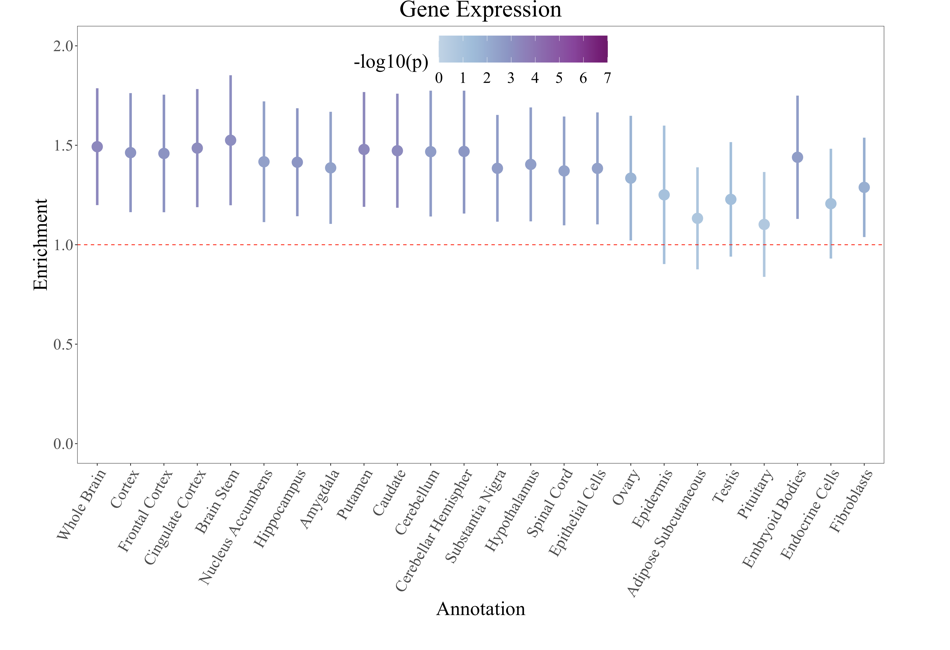

Figure S16c. Enrichment of Gene Expression Annotations for covariance between the *g*-factor and psychotic disorders factor. For comparative purposes, dots are depicted in descending order based on the point estimate for enrichment of the *g-*factor as per Figure S7c. Dots are shaded according to the significance of the enrichment estimate, with dots that were significant at a Bonferroni corrected threshold for 155 tests depicted with a *. The red dashed line reflects the null (enrichment = 1). Error bars depict 95% CIs. The scaling of the y-axis across enrichment graphs differs due to discrepant ranges in CIs across annotations.

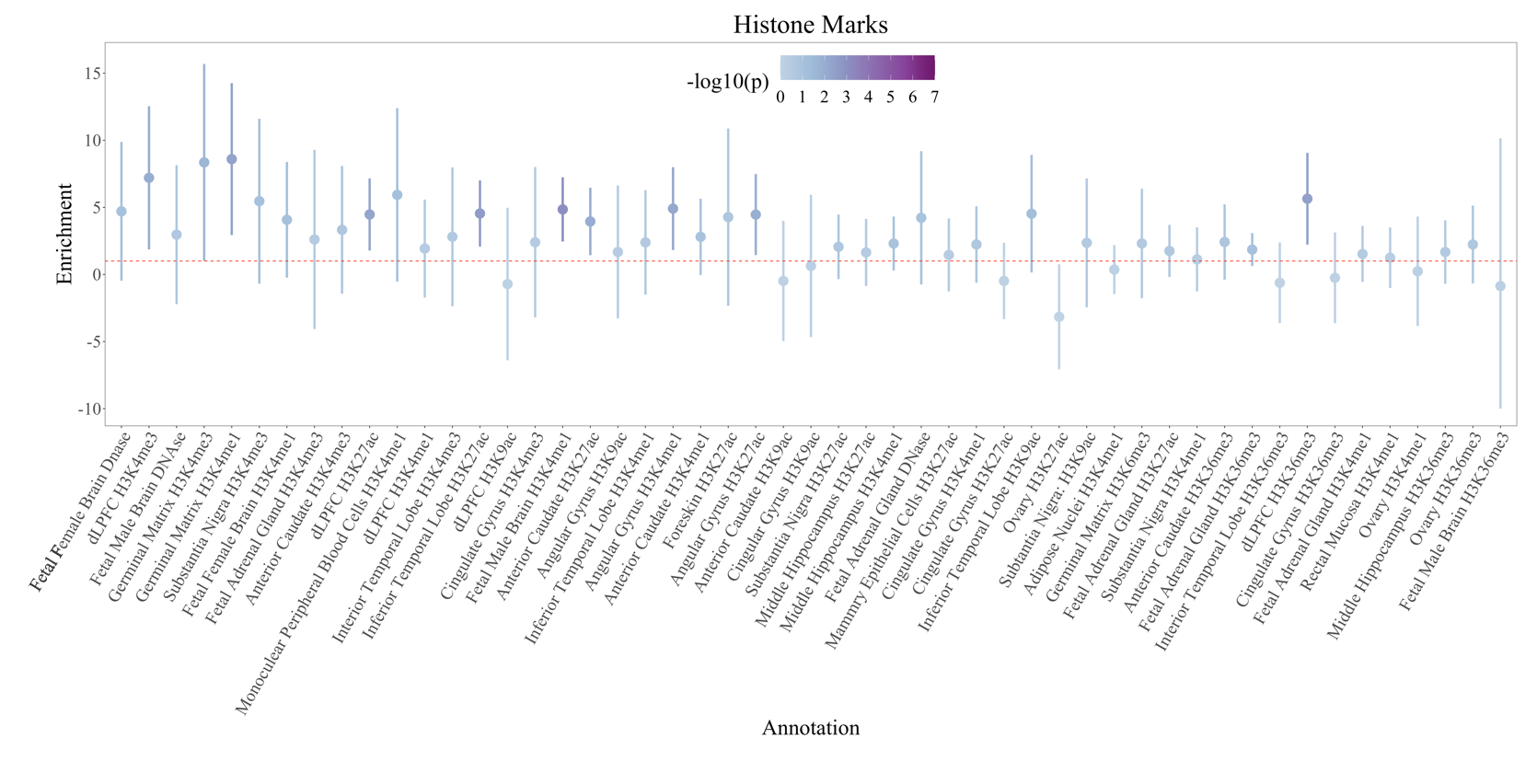

Figure S16d. Enrichment of Histone Mark Annotations for covariance between the *g*-factor and psychotic disorders factor. For comparative purposes, dots are depicted in descending order based on the point estimate for enrichment of the *g-*factor as per Figure S7d. Dots are shaded according to the significance of the enrichment estimate, with dots that were significant at a Bonferroni corrected threshold for 155 tests depicted with a *. The red dashed line reflects the null (enrichment = 1). Error bars depict 95% CIs. The scaling of the y-axis across enrichment graphs differs due to discrepant ranges in CIs across annotations.

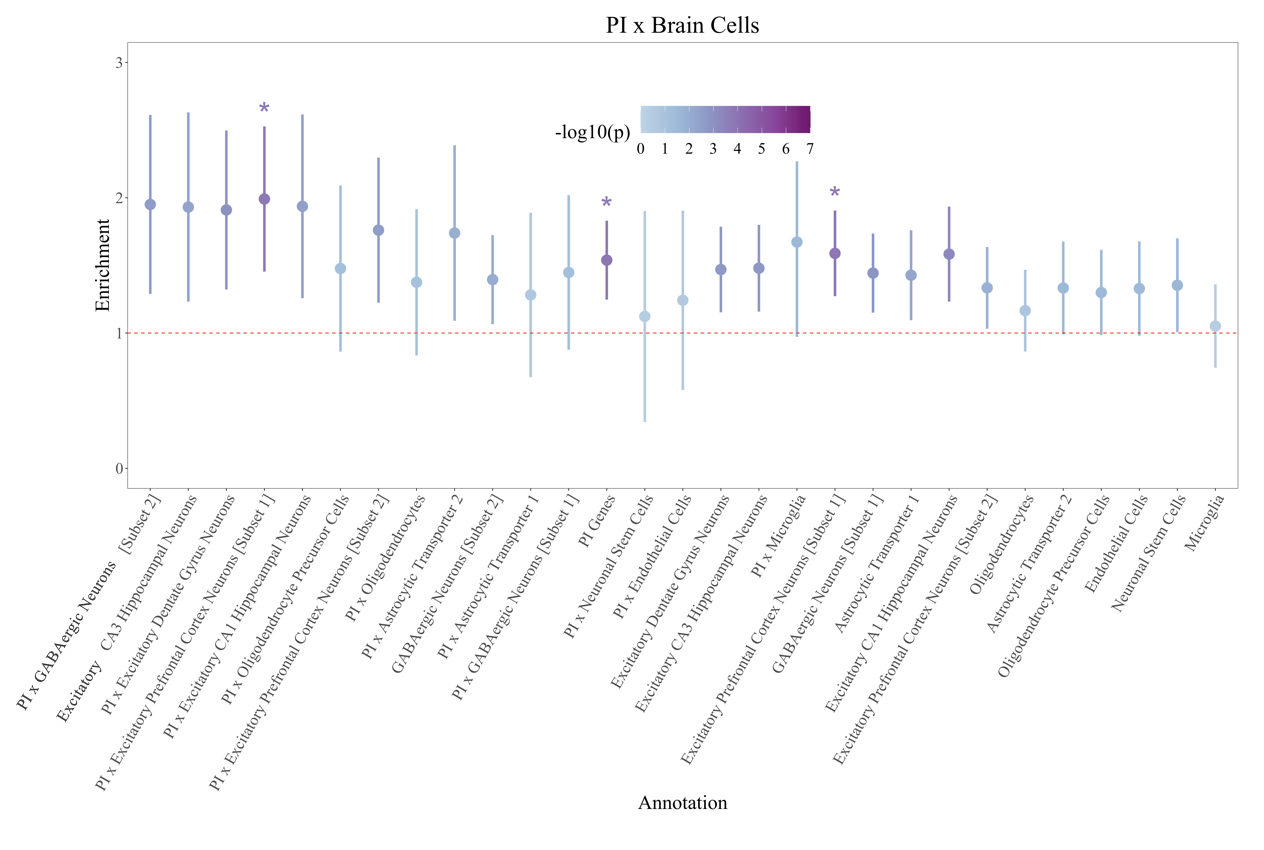

Figure S16e. Enrichment of PI x Brain Cell Annotations for covariance between the *g*-factor and psychotic disorders factor. For comparative purposes, dots are depicted in descending order based on the point estimate for enrichment of the *g-*factor as per Figure S7e. Dots are shaded according to the significance of the enrichment estimate. The red dashed line reflects the null (enrichment = 1), error bars depict 95% CIs, and dots that were significant at a Bonferroni corrected threshold for 155 tests are depicted with a *.
